# Redefining Non-Invasive Post-Transplant Surveillance: A Bayesian Meta-Analysis and Decision Curve Framework for Donor-Derived Cell-Free DNA in Heart Transplantation

**DOI:** 10.64898/2026.05.15.26353184

**Authors:** Jabez David John, Fathimathul Henna, Farah Waseem, Mohammad Aitzaz Hassan, Zaryab Bacha, Muhammad Mukhlis, Bilal Khan Mohammed, Shamikha Cheema, Kesha Shah

## Abstract

Donor-derived cell-free DNA (dd-cfDNA) is increasingly used for post transplantation non-invasive surveillance; however, its clinical interpretation remains inconsistent, with widely ranging thresholds and is typically applied as a single binary cutoff in literature. The optimal decision framework for rule-out and rule-in decisions, and whether a single threshold remains clinically meaningful, are currently uncertain. We performed a Bayesian hierarchical summary receiver operating characteristic (HSROC) meta-analysis of 14 studies (1,763 patients) evaluating dd-cfDNA against endomyocardial biopsy. To account for serial testing within individuals, we applied a cluster-corrected design effect, reducing 6,103 observations to 2,518 effective tests. Threshold-dependent sensitivity and specificity were modelled continuously. We compared a conventional single-threshold approach with a data-driven adaptive framework defining rule-out and rule-in thresholds and evaluated clinical utility by decision-curve analysis across rejection prevalences from 1% to 50%, incorporating repeat-testing strategies. The pooled area under the HSROC curve was 0.78 (95% CrI, 0.67-0.84). The Youden-optimal threshold (0.20%) yielded balanced sensitivity (0.77) and specificity (0.77) but failed to support clinical objectives of diagnosis. An adaptive framework identified a rule-out threshold of 0.16% (sensitivity 0.80) and a rule-in threshold of 0.48% (specificity 0.90), defining a indeterminate / grey zone. The residual one-in-five false-negative rate at the rule-out anchor reflects low-grade, non-cytolytic rejection, the imperfect histological reference standard and fractional suppression of the donor signal, rather than the statistical model; a result above the rule-in anchor carries a positive predictive value of approximately 38% at 10% prevalence and denotes an indication for tissue diagnosis and multimodal investigation, not for empiric treatment. Across low-to-intermediate prevalence, dd-cfDNA-guided strategies exceeded both the biopsy-all and monitor-all reference strategies; among testing strategies, repeat-if-borderline achieved the highest net benefit across the majority of the prevalence-threshold space and sustained positive net benefit over the widest operating range of any strategy, reducing false-positive biopsies without materially compromising detection. At high prevalence, where a first elevated result is usually true, biopsy-all became competitive. A single threshold is therefore clinically inadequate for post-transplant surveillance. Our tri-state, prevalence-aware framework integrating rule-out, indeterminate, and rule-in zones with selective repeat testing, more accurately reflects biomarker behavior and yields greater net benefit than any single cutoff because these anchors are pooled, population-level estimates rather than universal constants, programs should adopt this architecture and calibrate their own high-sensitivity rule-out and high-specificity rule-in thresholds to their local assay and population.

## Introduction

Post-heart transplant surveillance is mandatory, irrespective of the clinical symptoms exhibited by the patients, as rejections are frequently subclinical; approximately 12 % of heart transplant recipients experience moderate-to-severe rejection within a few years of transplant. (1–3) Although an endomyocardial biopsy (EMB) is invasive and often repeated, it remains the gold standard diagnostic tool. It typically functions as a binary switch in clinical decision-making, offering histological evidence that informs both the diagnosis and management of the patient. There is a clear unmet need to seek alternatives to EMB and promote non-invasive strategies, such as the use of blood biomarkers, gene expression profiles (GEP), and donor-derived cell-free DNA (dd-cfDNA). (4)

Donor-derived cell-free DNA (dd-cfDNA) is a promising diagnostic tool because of its ability to detect graft injury by measuring the elevation of donor DNA in the recipient’s bloodstream. The literature has consistently correlated dd-cfDNA with allograft injury (5–6). Clinical trials have demonstrated its potential (7), showing that it can detect transplant injury at an earlier stage, facilitate timely intervention, reduce the risk of full-blown rejection, and enable personalized immunosuppression, thereby potentially improving the long-term outcomes (8). Despite this strong biological rationale and growing clinical evidence, dd-cfDNA has yet to achieve standardized implementation in clinical practice, highlighting a gap between its demonstrated promise and widespread adoption.

As a continuous biomarker, its diagnostic meaning depends entirely on the threshold applied. The lack of standardized guidelines has resulted in optimal cutoffs across studies ranging from 0.1% to 0.55%, advocated by different centers and assays, posing a significant challenge. These discrepancies in threshold determinations hinder meaningful comparisons of results across studies and preclude the accurate estimation of dd-cfDNA’s true diagnostic performance, as pooling data from different thresholds conflates distinct operating points and obscures validity.

Prior meta-analyses pooled sensitivity and specificity across studies that used incompatible dd-cfDNA thresholds, conflating distinct operating points, and reported a single Youden-derived cutoff as a clinical recommendation (9). While the observation that a single dichotomous threshold is imperfect is not itself novel, no prior synthesis has modelled the threshold as a continuous covariate across the pooled evidence, derived explicit rule-out and rule-in anchors from the posterior, or validated the resulting decision structure on clinical net benefit. Those are the contributions of the present work.

We aimed to identify the optimal dd-cfDNA threshold for post-transplant rejection surveillance. We approached this study with the understanding that a test designed to rule out rejection requires high sensitivity, and a test designed to confirm rejection before proceeding to biopsy requires high specificity. These have different clinical purposes and may require different thresholds; if a single threshold can serve either or both, and whether any threshold, however derived, translates into actual clinical net benefit are gaps in the literature.

This Bayesian diagnostic test accuracy (DTA) meta-analysis aimed to identify whether a single optimal threshold is meaningful while providing a clinically grounded probabilistic decision framework to support clinicians in decision-making, regardless of the outcome.

## Methods

This DTA meta-analysis was conducted in accordance with the Preferred Reporting Items for Systematic Reviews and Meta-Analyses (i.e., PRISMA) (10) standards. This study was pre-registered with PROSPERO (CRD42025636627).

### Search Strategy

We conducted a diligent and computerized search across the PubMed, Medline, PubMed Central, Embase, Scopus, and Cochrane Central Registry databases from their inception. We recruited studies comparing dd-cfDNA and EMB in heart transplant recipients to evaluate graft rejection using in-depth search strings. The keywords were united using Medical Subject Headings (MeSH) terms.

### Selection

We systematically assessed the citations of the included articles and relevant review articles to identify further eligible studies based on a strict inclusion framework. The articles were uploaded to Rayyan. Duplicates were removed, and two authors independently screened and reviewed eligible articles. A senior author resolved discrepancies.

### Bias Assessment

QUADAS-2 (11) was used to assess the risk of bias and applicability of the included diagnostic accuracy studies

### Data Extraction

Data from the remaining articles were extracted and meticulously filed into a Microsoft Excel spreadsheet based on the *a priori* data extraction sheet. For the Bivariate diagnostic accuracy the true positives (TP), false positives (FP), false negatives (FN), true negatives (TN) and the physical dd-cfDNA threshold (%) applied in each study were extracted from the primary literature. Per-study sensitivity and specificity were calculated as

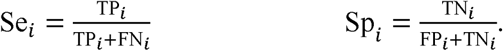

To stabilise the Markov chain Monte Carlo (MCMC) sampling, the threshold covariate was centred at the overall mean of all included study thresholds without altering the shape of the threshold-performance relationship.

### Correcting within-study clustering

The included studies contained multiple samples from the same patient were taken, and the data points are not independent, we deflated the count by design effect (DEff_i)

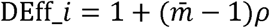

where *m̄* is the mean number of tests per patient and ρ is the intra-patient correlation. A conservative primary value of ρ = 0.50 was used for the cluster correction; robustness of the downstream decision-curve conclusions to the sequential-testing correlation was assessed by varying the Gaussian-copula correlation across ρ ∈ {0.30,0.50,0.70}.

### Bayesian HSROC Model

The corrected data was fit into a hierarchical summary receiver operating characteristic (HSROC) model where ddLcfDNA threshold was a covariate. The model estimates the HSROC curves across the thresholds accommodating between-study heterogeneity. Weakly informative priors were placed on all hyperparameters. Fitting used JAGS 4.3.2 with four MCMC chains (40,000 posterior draws). Convergence was confirmed with Gelman-Rubin diagnostics with (*R̂* < 1.01). A quadratic threshold term was tested however it did not improve the model’s fit (ΔDIC < 2).

### Model Evaluation

Predictive performance was assessed with Pareto-smoothed importance-sampling leave-one-out cross-validation (PSIS-LOO). When the Pareto-k diagnostic exceeded 0.7, we substituted an exact refit with proper marginalisation over study-specific random effects as recommended in modern Bayesian workflows (12)

### Summary Operating Points

Since the diagnostic accuracy of dd-cfDNA depends on the selected threshold, we decided to characterise its performance across the full range of clinically plausible cutoffs for which we constructed a continuous threshold-specific HSROC model and derived two complementary summary measures.

1. The entire threshold grid which provides a global estimate of sensitivity and specificity that does not presuppose any particular cutoff.
2. The model evaluates at the mean threshold observed among the included studies, permitting comparison with conventional meta-analytic summaries that assume an implicit common cutoff.

### Heterogeneity and Continuous Age Meta-Regression

Between-study heterogeneity was quantified from the joint posterior distributions of the HSROC random effects: the variance in test accuracy 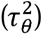 and the variance in the threshold effect 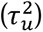. A continuous metaLregression was preferred over a binary subgroup analysis because Adult Congenital Heart Disease patients blur the paediatric/adult distinction, and ageLrelated physiological effects on ddLcfDNA are expected to act continuously rather than categorically. Instead, the study-level centred median age was introduced as a continuous linear covariate in the location parameter (*λ_i_*) of the HSROC model. A weakly informative Gaussian prior (*β_age_* ∼ N(0,0.25)) was used, and the influence of age was assessed by checking whether the 95% credible interval for the age coefficient included zero.

### Assessment of Publication Bias

Publication bias was assessed with Deeks’ funnel plot asymmetry test. Egger’s test and standard funnel plots are unsuitable for diagnostic test accuracy meta-analyses because of the artefactual correlation between diagnostic odds ratios and their variances. A linear regression of the log diagnostic odds ratio on the inverse root of the effective sample size was therefore performed. A Haldane-Anscombe continuity correction (+0.5) was applied to zero-cell studies to prevent infinite variance limits. Following standard DTA methodology, a significance threshold of α = 0.10 was used to flag meaningful asymmetry.

### Analytical Framework: Classification versus Risk Quantification

Two distinct statistical traditions govern the evaluation of a diagnostic test, and the distinction between them determines which metrics are admissible for which question. The first, classification, asks whether a test can discriminate between the diseased and non-diseased states. Its natural metrics are sensitivity, specificity, the diagnostic odds ratio, the area under the receiver operating characteristic curve, and summary indices such as the Youden index. These quantities are properties of the test itself: they are estimated within the diseased and non-diseased strata separately and are therefore, by construction, invariant to disease prevalence. The question they answer is whether the test works.

The second tradition, risk quantification, asks how a test already shown to work should be deployed to alter clinical action. Its natural metrics are the post-test probability, the calibration of predicted risk, and decision-analytic quantities such as net benefit. These are not properties of the test in isolation but of the test embedded within a population and a decision. They depend explicitly on prevalence and on the relative weight the clinician assigns to a missed rejection episode against an unnecessary biopsy. The question they answer is how a given result should trigger an invasive procedure in practice.

We report both, deliberately and in sequence. We first computed the Youden index because it is the appropriate and statistically sound answer to the first question. The Youden index identifies the cutoff maximising the sum of sensitivity and specificity, weights the two error types equally, and requires no assumption regarding prevalence or the relative cost of either error. Under those assumptions the Youden-derived cutoff, including the 0.20% value derived here and the comparable values reported across the prior literature, is a valid and internally consistent summary of the intrinsic discriminatory capacity of the biomarker. It establishes that dd-cfDNA carries genuine diagnostic information about allograft rejection, and we regard the foundational studies that derived such thresholds as having answered their intended question correctly and by appropriate means.

The constraint on the Youden index is therefore one of scope rather than of statistical validity. Precisely because it is prevalence-invariant and cost-symmetric, it cannot by construction answer the second question. Post-transplant surveillance is neither prevalence-invariant, since per-test rejection prevalence across the included studies spans approximately 0.8% to 52%, nor cost-symmetric, since an unnecessary endomyocardial biopsy and a missed rejection episode are not equivalent harms. Once the question shifts from whether dd-cfDNA discriminates to when a dd-cfDNA result should commit a patient to an invasive procedure, a prevalence-aware and cost-weighted metric becomes mandatory rather than discretionary.

Decision curve analysis supplies exactly this. Net benefit places sensitivity and specificity on a common utility scale defined by the threshold probability, the probability of rejection at which a clinician is indifferent between proceeding to biopsy and withholding it, and evaluates the resulting trade-off at the prevalence actually obtaining in the population of interest.

The analyses that follow should accordingly be read in that order and with that division of labour: the Youden analysis as a classification benchmark that validates the biomarker, and the decision curve analysis as a separate, decision-theoretic evaluation that determines how the validated biomarker should be operationalised. Our argument throughout is directed at the practice of importing a classification-derived cutoff directly into a clinical decision rule without re-evaluating it under a decision-theoretic loss function, and not at the derivation of that cutoff itself.

### Optimal Threshold Analysis

#### Youden index

To identify the single threshold that mathematically maximises diagnostic performance, we computed the Youden index (U) = sensitivity + specificity -1 at every point on the threshold grid. The cutoff that maximised the posterior mean of (U) was taken as the optimal threshold in the classical test-evaluation sense.

#### Adaptive sensitivity anchor

We sought the highest achievable sensitivity that could serve as a rule-out criterion. Because a threshold providing near-perfect sensitivity would be clinically desirable, we began with a target of 95L% sensitivity and examined whether at least 50L% of posterior draws could sustain that sensitivity at any cutoff. If the target was unattainable, we lowered it in 5-percentage-point decrements until we reached a level supported by the majority of posterior draws. An analogous procedure was followed for specificity, starting at a target of 95L% and adaptively lowering by 5-percentage-point it if necessary. This adaptive walkLdown procedure ensures that the reported anchors are dataLdriven. Instead of imposing an arbitrary sensitivity or specificity level that the literature cannot support, we allowed the evidence to determine what level is realistically attainable and report the corresponding cutoffs.

### Decision Curve analysis

To evaluate whether the mathematically optimal threshold translates into clinically optimal decisions, we compared five surveillance strategies across the full clinically relevant range of rejection prevalence (1-50%), rather than at a small number of fixed values. This range spans true asymptomatic surveillance through for-cause and DSA-positive antibody-mediated rejection populations and is supported by the per-test prevalence observed across the included studies (∼0.8% to 52%). Net benefit was evaluated over a two-dimensional grid of rejection prevalence (1-50%) and biopsy threshold probability, and summarized both as the region in which each strategy is the best available choice and as each strategy’s absolute net benefit.

The five strategies were as follows:

1. Biopsy all where endomyocardial biopsy performed on every patient, representing the current standard in many centres.
2. Monitor all strategy were clinical surveillance without biopsy, representing zero intervention.
3. Single test at the Youden cutoff where the biopsy triggered when dd-cfDNA crosses the Youden-optimal threshold; this strategy should be optimal if that threshold captures the best sensitivity-specificity trade-off, and it serves as the reference point for single-test decisions.
4. Repeat-if-negative is when the first test falls below the Youden cutoff, retesting is performed after 3-7 days, with biopsy reserved for patients whose second test is also positive; this strategy aims to reduce unnecessary biopsies in patients with transiently low dd-cfDNA who nonetheless harbour rejection.
5. Repeat-if-borderline is where the adaptive sensitivity and specificity anchors are used as rule-out (TL) and rule-in (TL) thresholds; results in the intermediate zone (TL-TL) are repeated, and biopsy is performed only if the repeat remains intermediate or high; this approach mirrors clinical practice, where borderline results typically prompt retesting before committing to an invasive procedure.

For strategies involving two sequential tests, joint probabilities of the binary outcomes were computed with a Gaussian copula (intra-patient correlation = 0.50), guaranteeing that all probabilities remain within [0,1]. Net benefit was calculated as

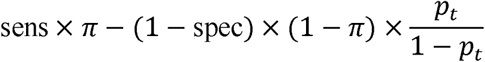

where *π* is the prevalence of rejection and *p_t_* is the threshold probability of biopsy. Posterior medians and 95% credible intervals of net benefit were generated across the threshold probabilities.

For sequential-testing strategies, the primary copula correlation was *ρ* ∈ {0.50}., with sensitivity analysis across *ρ* ∈ {0.30,0.50,0.70}.

### Software

The entire analysis was performed on R 4.5.0, with core modelling in JAGS via rjags, PSIS-LOO via loo, copula calculations via pbivnorm, and graphics via ggplot2.

Full search strings, study inclusion criteria, QUADAS-2 results, and detailed justification for all statistical modeling decisions are provided in the Supplementary Material.

## Results

### 1. Study characteristics and data summary (PRISMA diagram and DEff table)

We utilized specialized detailed search strategies across PubMed, Medline, PubMed Central, Scopus, Cochrane Central Registry, and Embase databases to identify 4,286 articles. After removing 253 duplicates, the remaining articles were rigorously screened. Following screening, the articles were assessed for eligibility, which led to the inclusion of 14 studies in the final analysis. Our final analysis encompassed 10 prospective (71.4%) and four retrospective (28.6 %) studies. The range of the different thresholds for diagnosing heart rejection using dd-cfDNA is 0.1-0.55.

Nine studies were conducted in the United States, two in Italy, and one each in Spain, Sweden, and Germany. The years of publication were extended from 2014 to 2024. The total number of patients included in the studies was 1763, and the percentage of women across the studies ranged from 25.5% to 70%. Tables 1 and 2 summarize the baseline characteristics of the included studies.

**Table 1:**
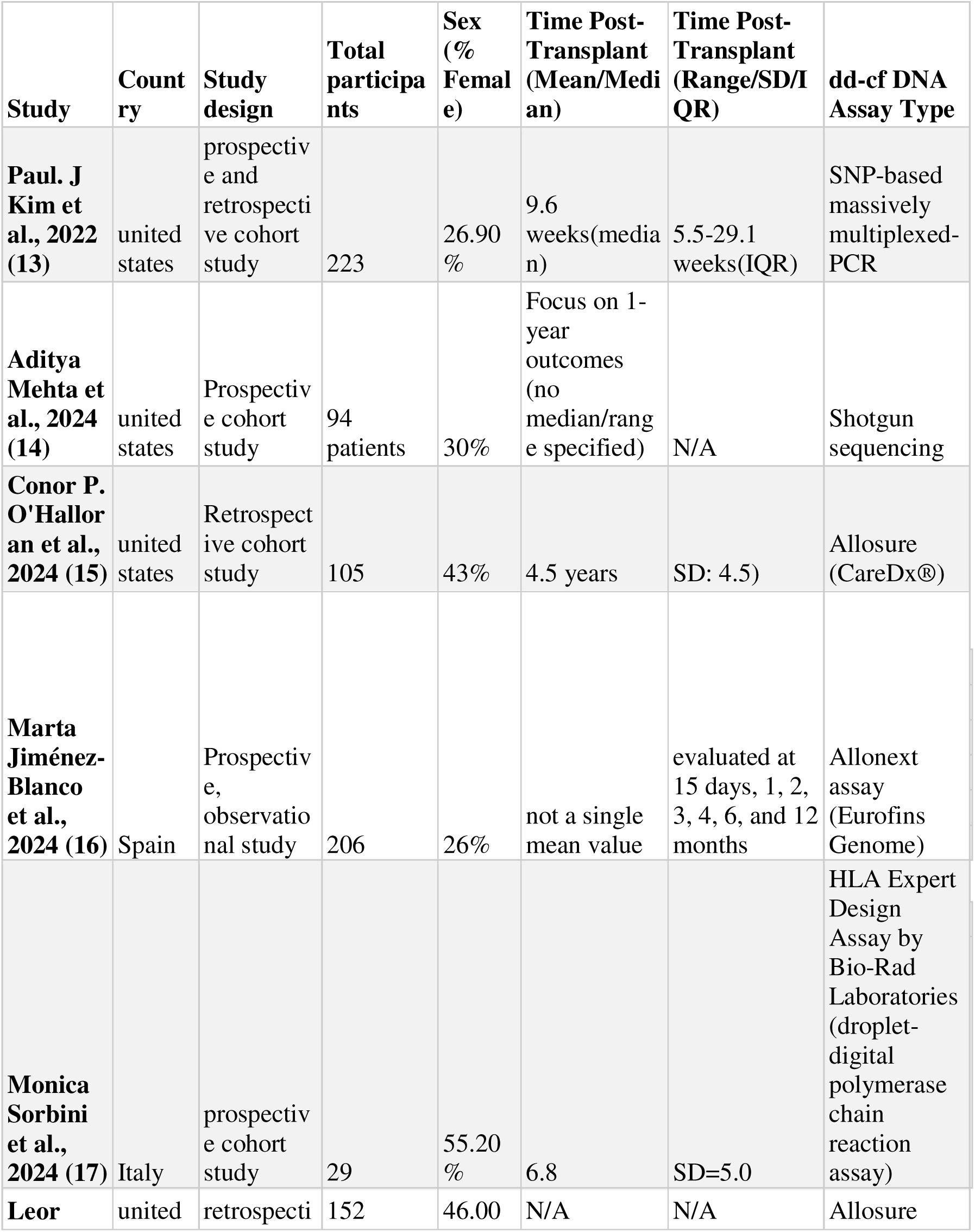
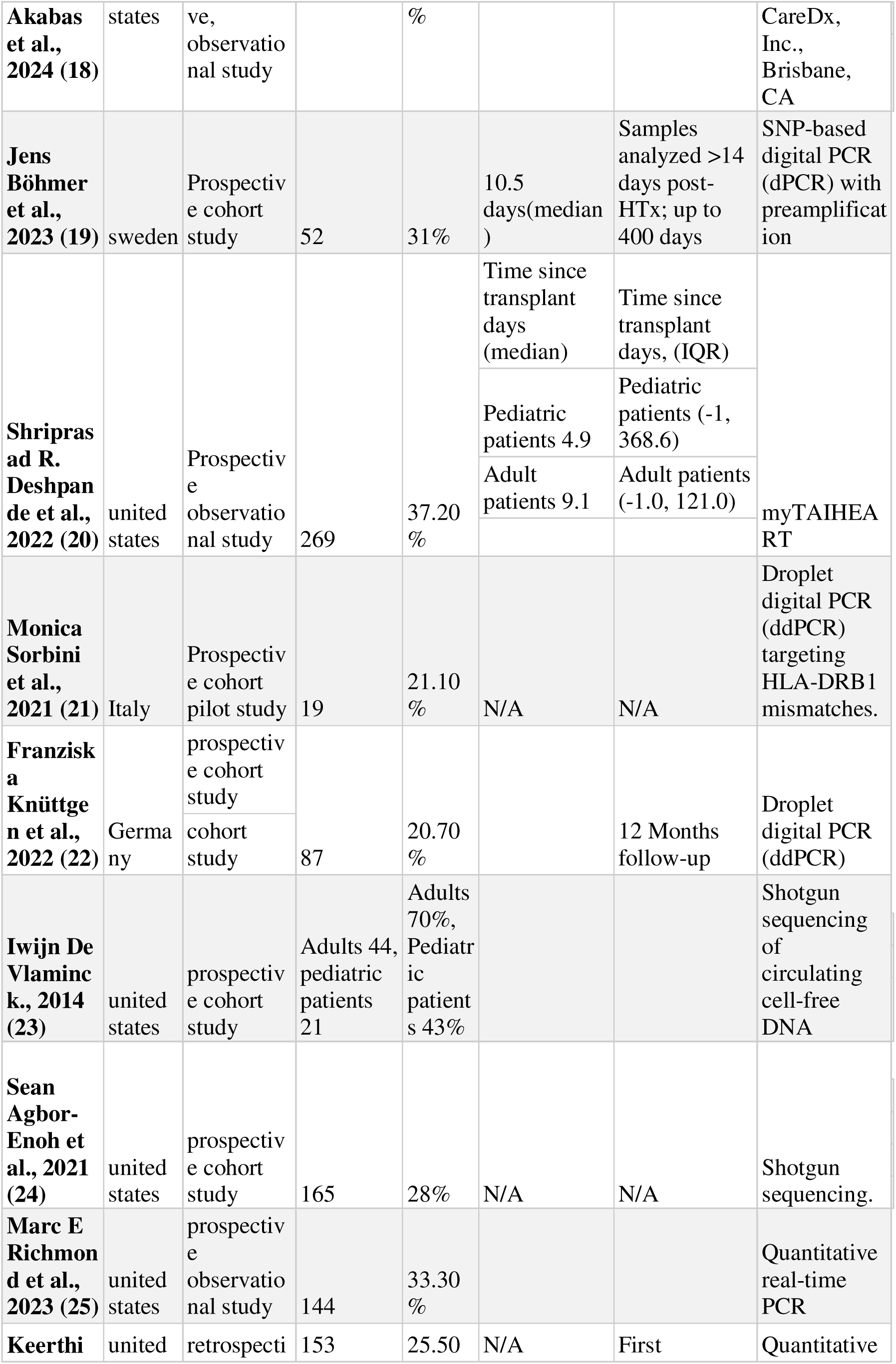
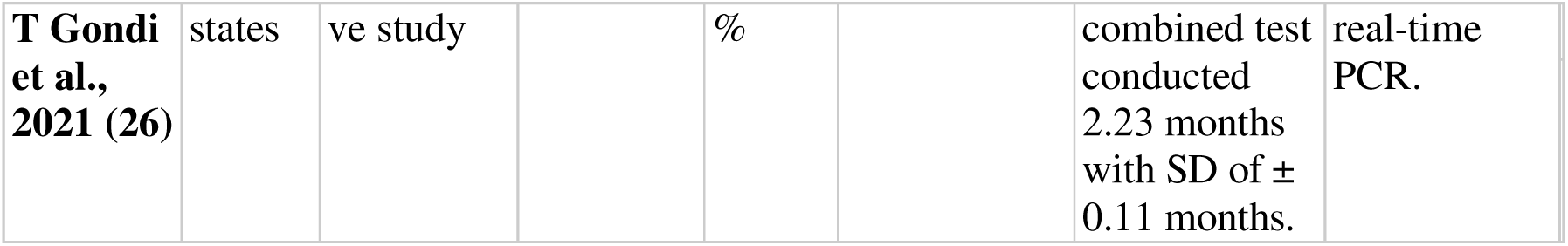
Study and Population Characteristics.

| Study | Country | Study design | Total participants | Sex (% Female) | Time Post-Transplant (Mean/Median) | Time Post-Transplant (Range/SD/IQR) | dd-cf DNA Assay Type |
| --- | --- | --- | --- | --- | --- | --- | --- |
| Paul. J Kim et al., 2022 (13) | united states | prospective and retrospective cohort study | 223 | 26.90 % | 9.6 weeks(median) | 5.5-29.1 weeks(IQR) | SNP-based massively multiplexed-PCR |
| Aditya Mehta et al., 2024 (14) | united states | Prospective cohort study | 94 patients | 30% | Focus on 1-year outcomes (no median/range specified) | N/A | Shotgun sequencing |
| Conor P. O'Halloran et al., 2024 (15) | united states | Retrospective cohort study | 105 | 43% | 4.5 years | SD: 4.5) | Allosure (CareDx®) |
| Marta Jiménez-Blanco et al., 2024 (16) | Spain | Prospective, observational study | 206 | 26% | not a single mean value | evaluated at 15 days, 1, 2, 3, 4, 6, and 12 months | Allonext assay (Eurofins Genome) |
| Monica Sorbini et al., 2024 (17) | Italy | prospective cohort study | 29 | 55.20 % | 6.8 | SD=5.0 | HLA Expert Design Assay by Bio-Rad Laboratories (droplet-digital polymerase chain reaction assay) |
| Leor | united | retrospective | 152 | 46.00 | N/A | N/A | Allosure |
| **Akabas et al., 2024 (18)** | states | ve, observational study |  | % |  |  | CareDx, Inc., Brisbane, CA |
| **Jens Böhmer et al., 2023 (19)** | sweden | Prospective cohort study | 52 | 31% | 10.5 days(median ) | Samples analyzed >14 days post-HTx; up to 400 days | SNP-based digital PCR (dPCR) with preamplification |
| **Shripras ad R. Deshpande et al., 2022 (20)** | united states | Prospective observational study | 269 | 37.20 % | Time since transplant days (median) | Time since transplant days, (IQR) | myTAIHEART |
|  |  |  |  |  | Pediatric patients 4.9 | Pediatric patients (-1, 368.6) |  |
|  |  |  |  |  | Adult patients 9.1 | Adult patients (-1.0, 121.0) |  |
| **Monica Sorbini et al., 2021 (21)** | Italy | Prospective cohort pilot study | 19 | 21.10 % | N/A | N/A | Droplet digital PCR (ddPCR) targeting HLA-DRB1 mismatches. |
| **Franziska Knüttgen et al., 2022 (22)** | Germany | prospective cohort study | 87 | 20.70 % |  | 12 Months follow-up | Droplet digital PCR (ddPCR) |
|  |  | cohort study |  |  |  |  |  |
| **Iwijn De Vlaminck, 2014 (23)** | united states | prospective cohort study | Adults 44, pediatric patients 21 | Adults 70%, Pediatric patients 43% |  |  | Shotgun sequencing of circulating cell-free DNA |
| **Sean Agbor-Enoh et al., 2021 (24)** | united states | prospective cohort study | 165 | 28% | N/A | N/A | Shotgun sequencing. |
| **Marc E Richmond et al., 2023 (25)** | united states | prospective observational study | 144 | 33.30 % |  |  | Quantitative real-time PCR |
| **Keerthi** | united | retrospective | 153 | 25.50 | N/A | First | Quantitative |
| **T Gondi et al., 2021 (26)** | states | ve study | | % | | combined test conducted 2.23 months with SD of ± 0.11 months. | real-time PCR. |

**Table 2:**
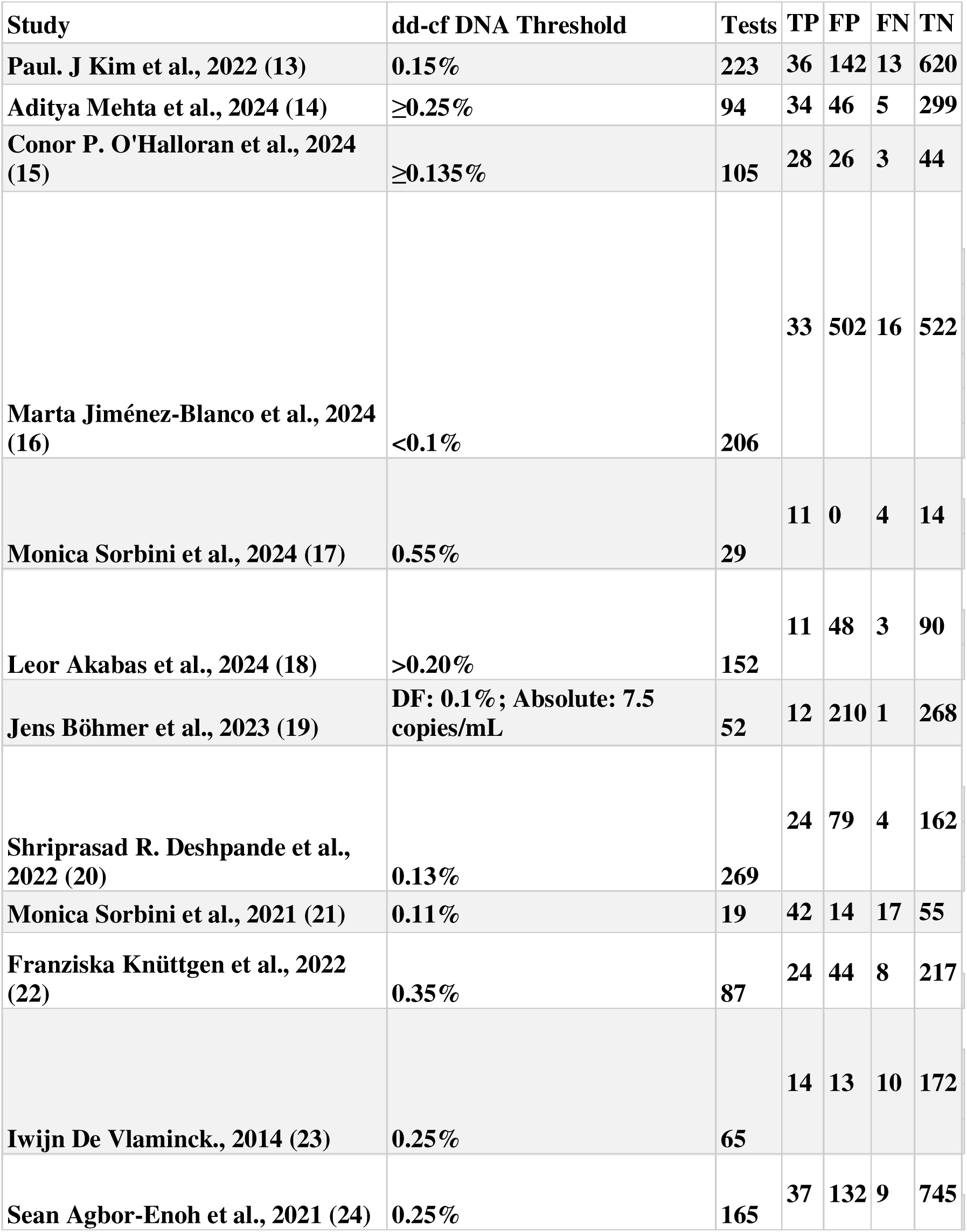
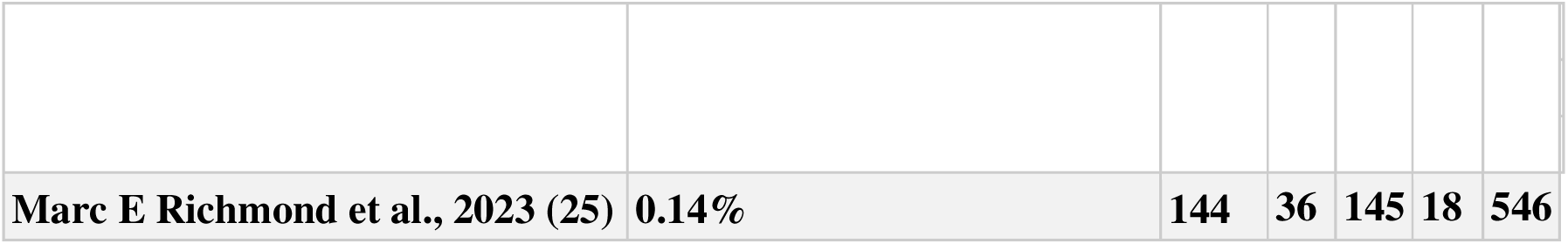
Diagnostic Data and Threshold Details.

Studies varied substantially in their tests-per-patient ratio (mean tests per patient, m□, range: 1.0-9.4), reflecting differences in sampling design. To avoid over-weighting studies with dense repeated sampling, effective sample sizes were computed using a cluster-corrected design effect, with ρ = 0.50 as the primary analysis. Under this assumption, the 6,103 reported tests corresponded to 2,518 effective tests (mean DEff = 2.42; per-study DEff range: 0.98-5.22).

### 2. HSROC model and AUC

The Bayesian HSROC model with threshold as a study-level covariate yielded a pooled AUC of 0.781 (95% CrI: 0.668-0.840), indicating a moderate-to-good discriminative ability of dd-cfDNA for acute rejection after heart transplantation. (Figure 1).

**Figure 1.**
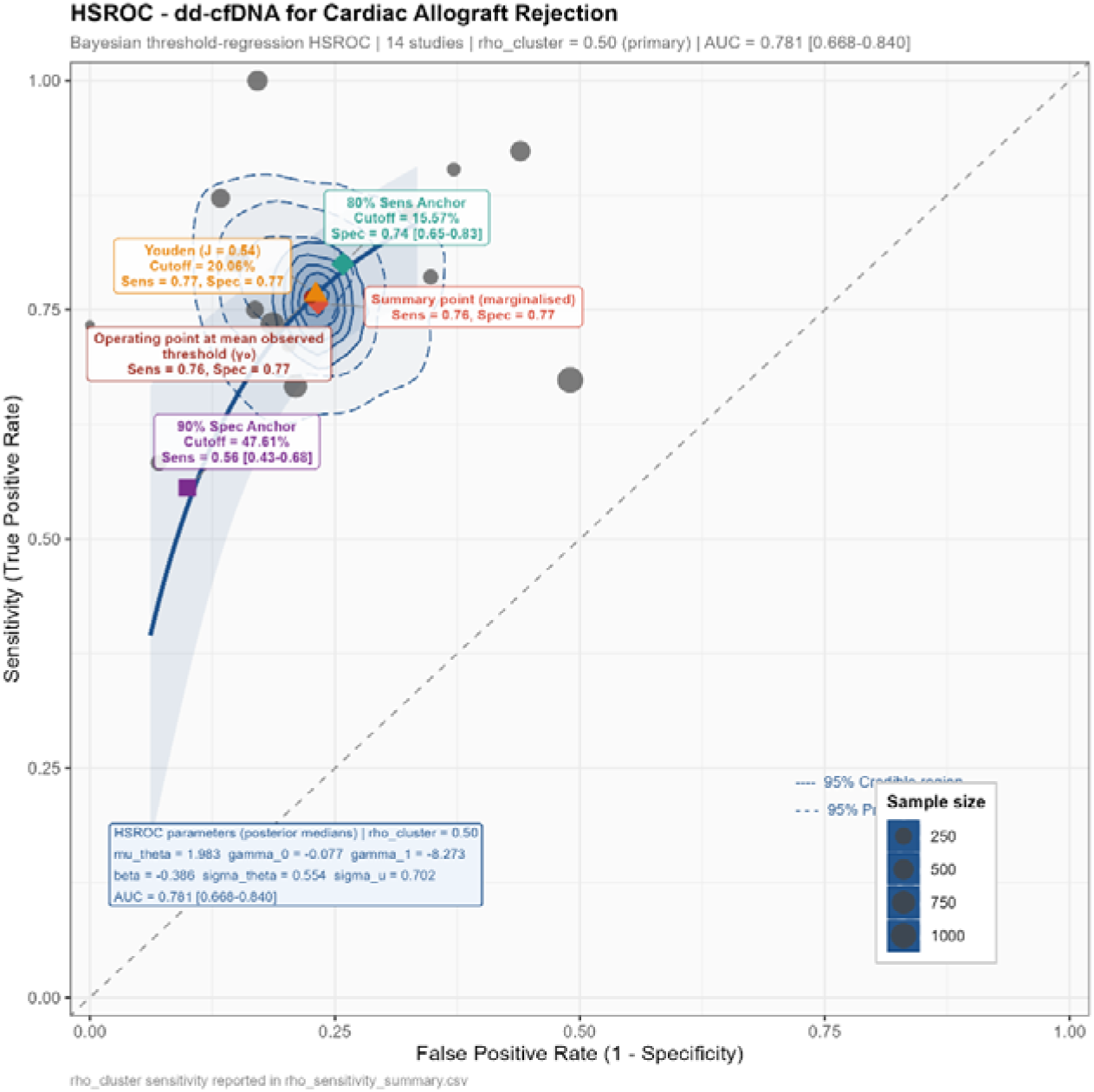
Bayesian hierarchical summary receiver operating characteristic (HSROC) curve for dd-cfDNA in detecting acute rejection after heart transplantation. The SROC curve (solid line) represents the posterior predictive relationship between sensitivity and specificity across the range of clinically studied dd-cfDNA thresholds (0.10%-0.55%). The shaded region denotes the 95% credible interval. The pooled area under the HSROC curve (AUC) was 0.781 (95% CrI: 0.668-0.840). The summary operating point at the mean observed threshold (γ□) is indicated. Individual study thresholds are plotted as points, sized proportionally to effective sample size after cluster correction (ρ = 0.50). DD-cfDNA, donor-derived cell-free DNA; HSROC, hierarchical summary receiver operating characteristic; CrI, credible interval.

Marginalizing posterior sensitivity and specificity over the empirical distribution of study-reported thresholds by the canonical HSROC summary point showcased a pooled sensitivity of 0.757 (95% CrI: 0.678-0.823) and specificity of 0.767 (95% CrI: 0.713-0.814), and at the mean observed threshold (γ), the operating point was sensitivity 0.764 (0.683-0.831) and specificity 0.775 (0.720-0.822). Detailed posterior estimates are presented in Table 3.

**Table 3.**
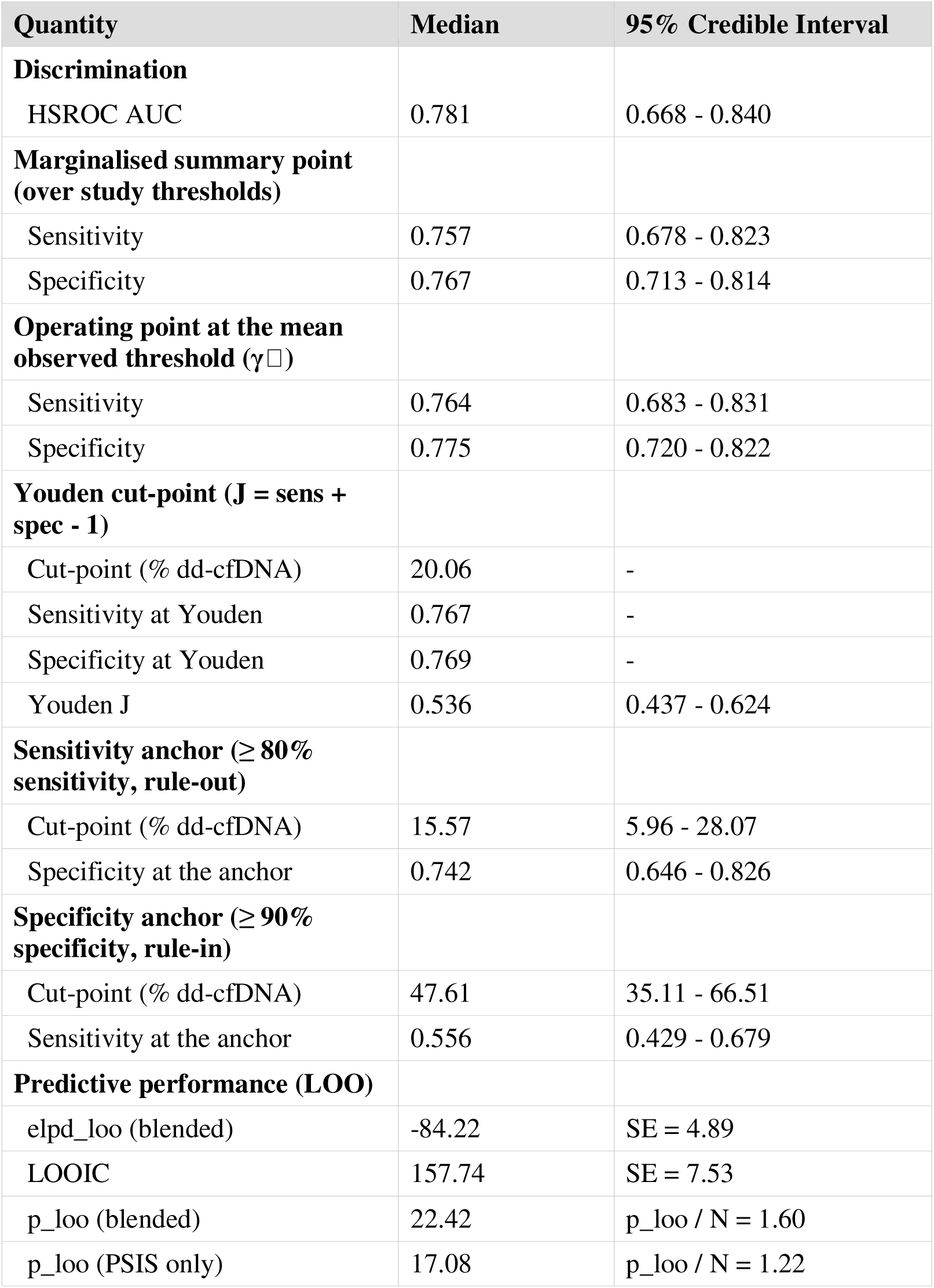
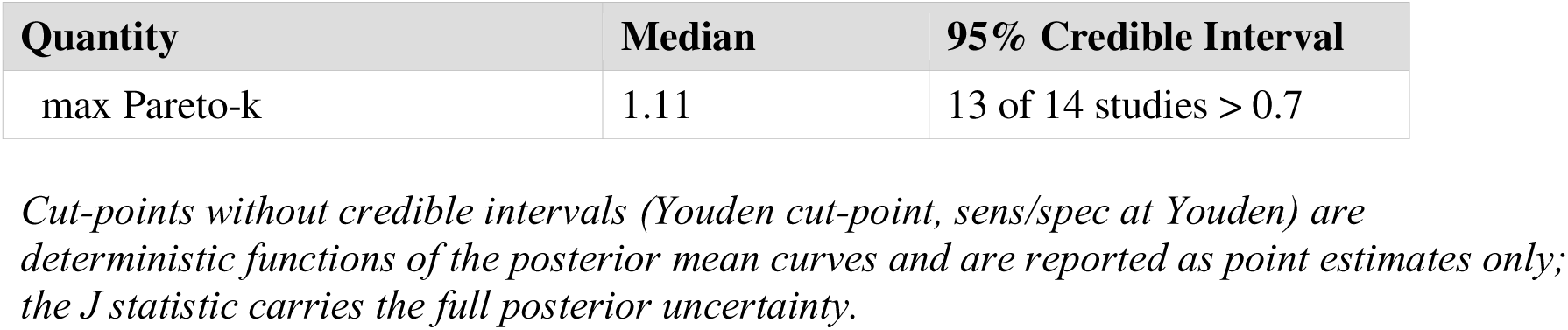
Primary results at ρ = 0.50. Posterior medians with 95% credible intervals.

### Youden optimal threshold

Posterior sensitivity and specificity as continuous functions of the dd-cfDNA threshold are shown in Figure 2. The data-driven Youden optimum corresponded to a dd-cfDNA cutoff of 0.20% (sensitivity, 0.767; specificity, 0.769; Youden’s J, 0.536; 95% confidence interval [CrI], 0.437-0.624).

**Figure 2.**
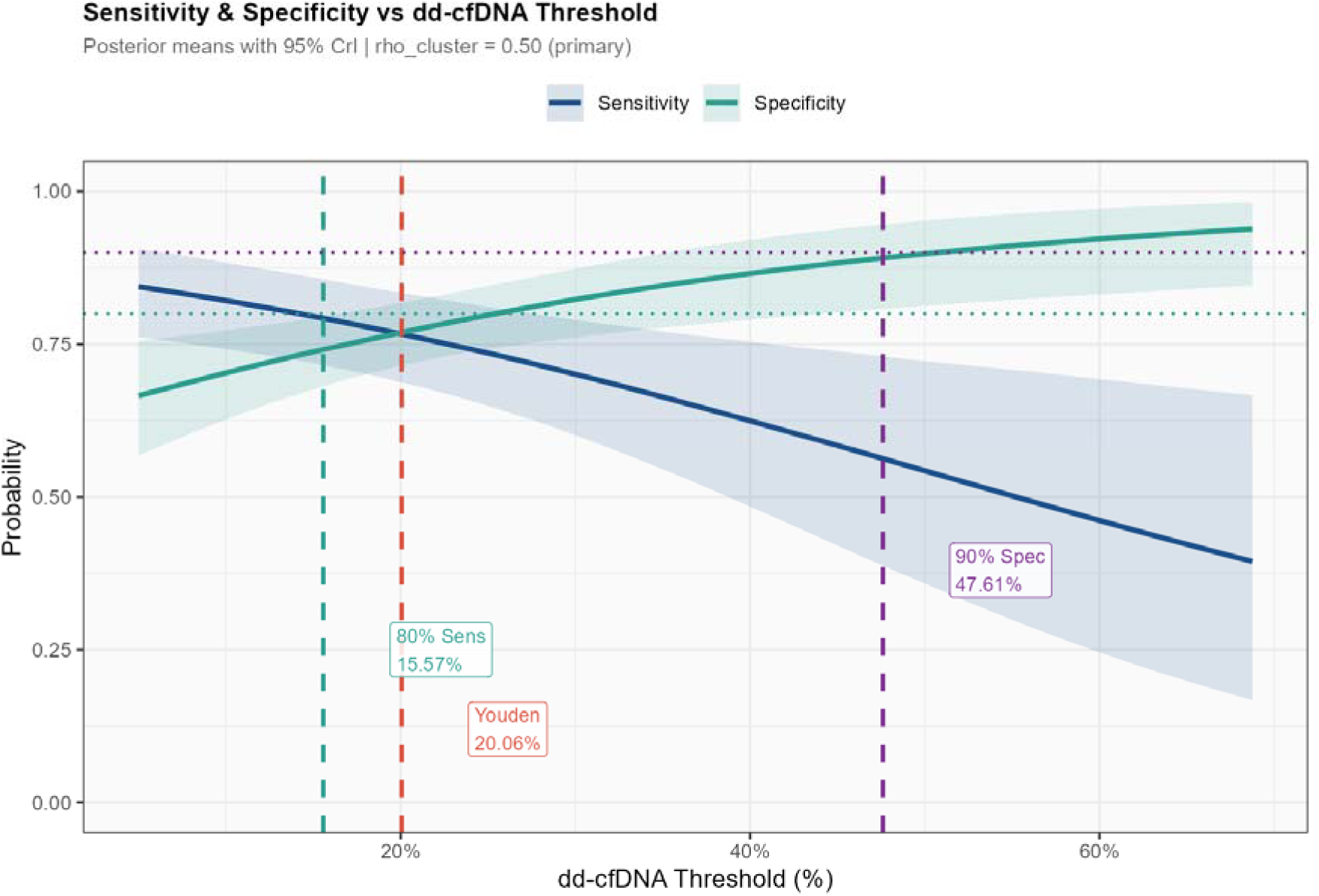
Posterior sensitivity and specificity as continuous functions of the dd-cfDNA threshold. Solid curves represent posterior median sensitivity (blue) and specificity (red) plotted across the full range of clinically plausible dd-cfDNA cutoffs (0.10%-0.55%). Shaded ribbons represent 95% credible intervals. The Youden-optimal threshold (0.20%) is indicated by a vertical dashed line, corresponding to a sensitivity of 0.767 and specificity of 0.769 (Youden’s J = 0.536; 95% CrI: 0.437-0.624). The rule-out anchor (T□ = 0.16%, sensitivity 0.80) and rule-in anchor (T□ = 0.48%, specificity 0.90) are indicated by vertical dotted lines with shading demarcating the three clinical zones. DD-cfDNA, donor-derived cell-free DNA; CrI, credible interval.

### 4. Forest plots

Forest plots of study-specific sensitivity (Figure 3) and specificity (Figure 4) showed substantial between-study variation, consistent with the heterogeneity in reported thresholds and study designs. Sensitivity estimates ranged across the included studies, with the pooled marginal summary (Figure 3) anchored at 0.757; specificity showed a similarly wide spread around the pooled summary of 0.767 (Figure 4).

**Figure 3.**
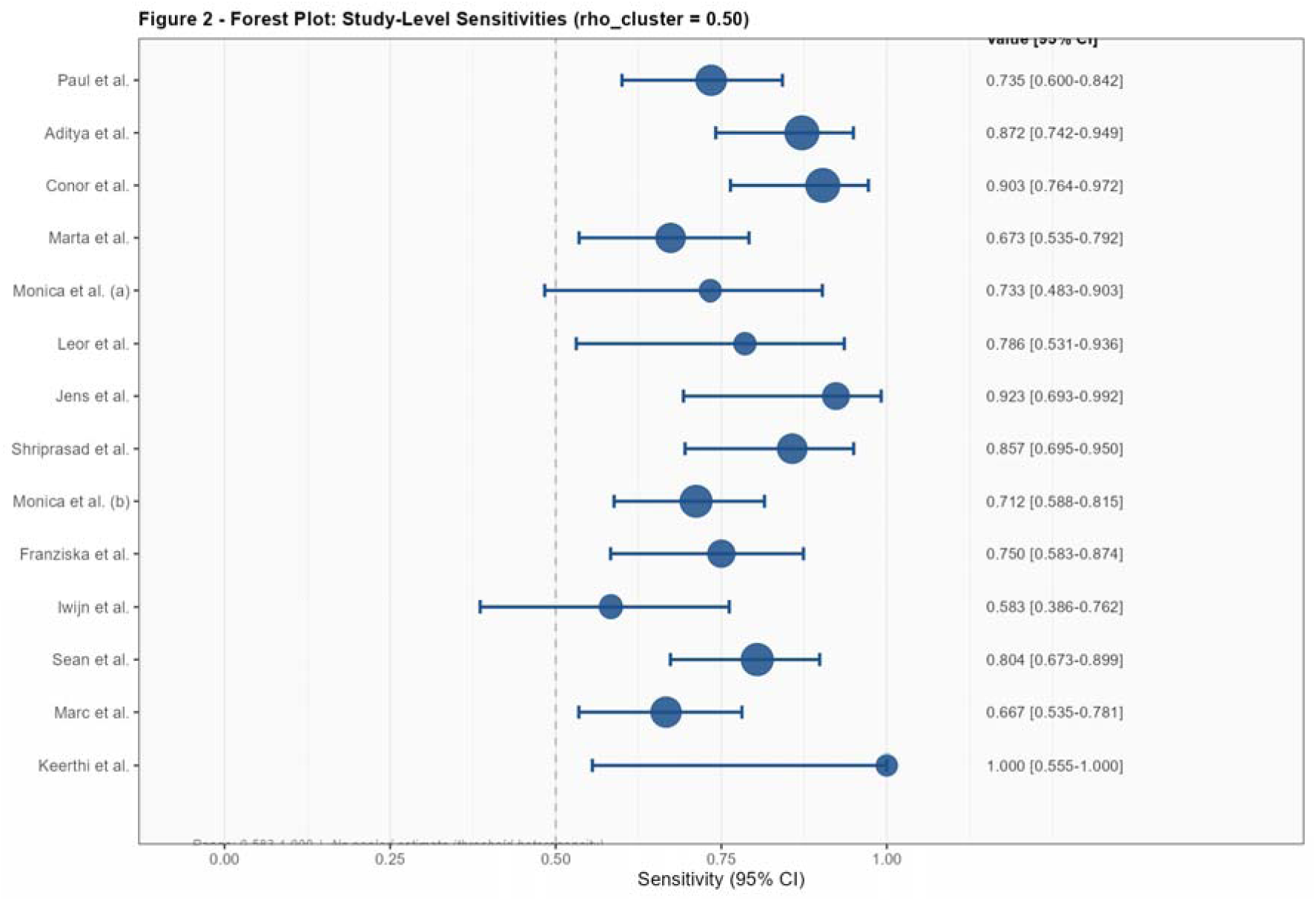
Forest plot of study-level sensitivity estimates for dd-cfDNA in detecting acute cardiac allograft rejection. Each row represents one of the 14 included studies. Point estimates (squares) are sized proportionally to effective weight after cluster correction (design effect, ρ = 0.50). Horizontal bars represent 95% confidence intervals. The pooled marginal sensitivity (0.757; 95% CrI: 0.678-0.823) is shown as a diamond at the bottom. Between-study variance in accuracy (τ²θ = 0.306; 95% CrI: 0.109-0.969) reflects substantial heterogeneity attributable to threshold variability across studies. DD-cfDNA, donor-derived cell-free DNA; CrI, credible interval.

**Figure 4.**
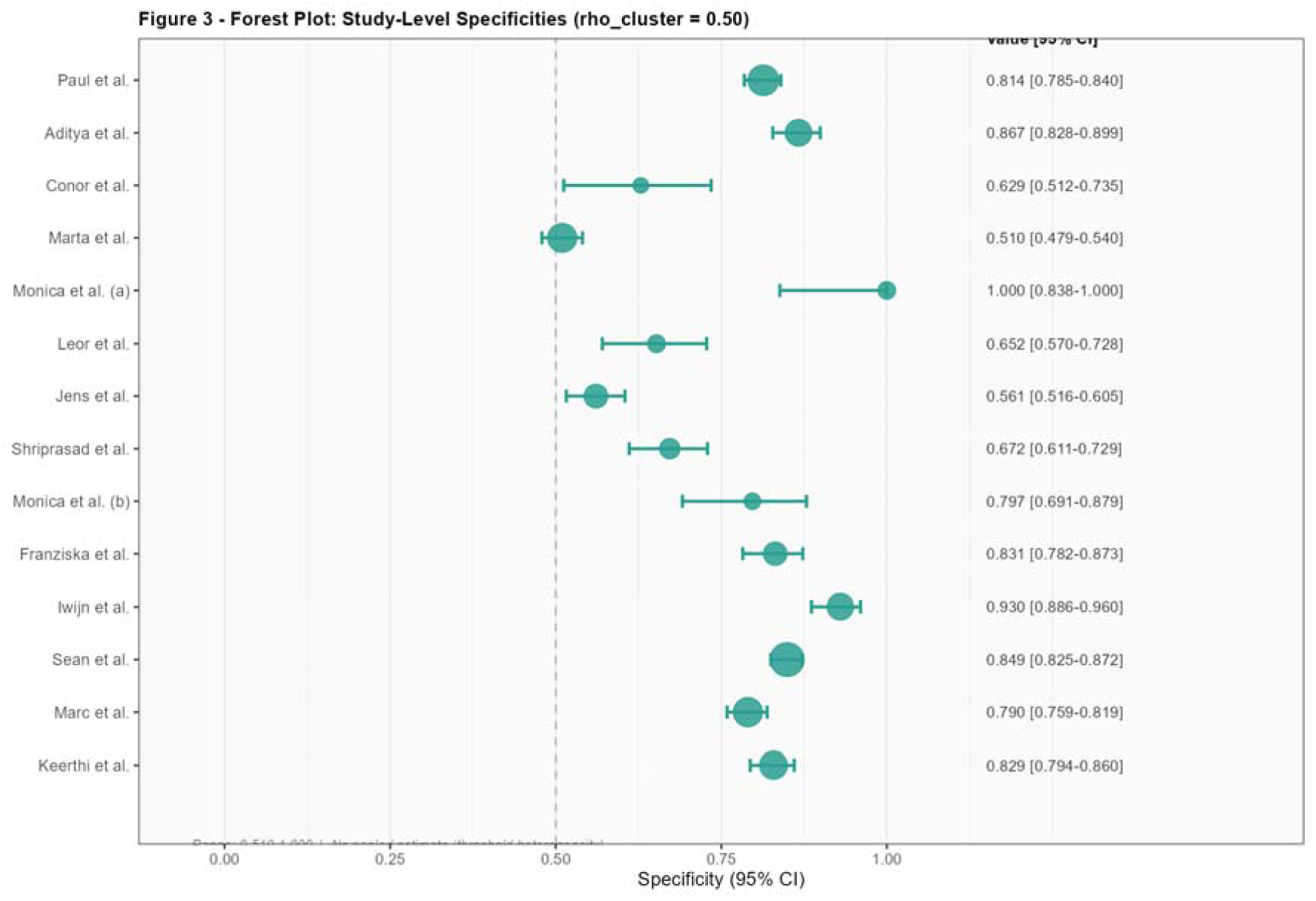
Forest plot of study-level specificity estimates for dd-cfDNA in detecting acute cardiac allograft rejection. Each row represents one of the 14 included studies. Point estimates (squares) are sized proportionally to effective weight after cluster correction (design effect, ρ = 0.50). Horizontal bars represent 95% confidence intervals. The pooled marginal specificity (0.767; 95% CrI: 0.713-0.814) is shown as a diamond at the bottom. Wide between-study spread is consistent with the substantial threshold variance (τ²u = 0.493; 95% CrI: 0.140-1.801) across the included studies. DD-cfDNA, donor-derived cell-free DNA; CrI, credible interval.

### 5. Adaptive sensitivity and specificity anchors

At a sensitivity target of 80%, at least 50% of the posterior draws supported this level of performance, yielding a required dd-cfDNA threshold of 0.156% (95% CrI: 0.060-0.281%) with a corresponding specificity of 0.742 (95% CrI: 0.646-0.826).

At a specificity target of 90%, at least 50% of posterior draws supported this level of performance, yielding a required dd-cfDNA threshold of 0.476% (95% CrI: 0.351-0.665%) with a corresponding sensitivity of 0.556 (95% CrI: 0.429-0.679).

Both anchor thresholds fell within the empirically studied range of reported dd-cfDNA cutoffs (0.10-0.55%), indicating that the posterior estimates remained clinically grounded. (Table 4,Figure 5)

**Figure 5.**
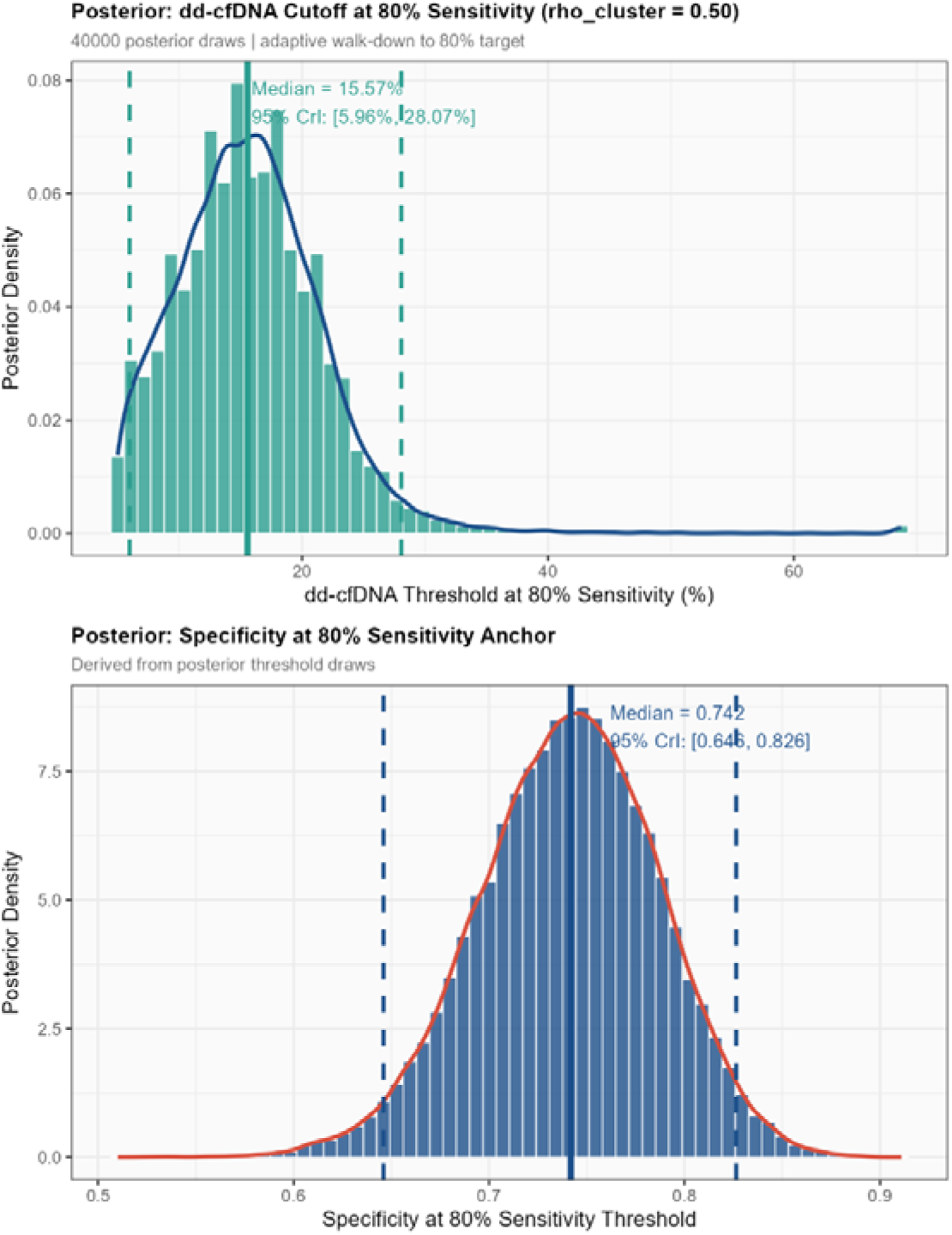
Posterior sensitivity and specificity as continuous functions of the dd-cfDNA threshold, with adaptive rule-out and rule-in anchors. Solid curves represent posterior median sensitivity (blue) and specificity (red) plotted across the full range of clinically plausible dd-cfDNA cutoffs (0.10%-0.55%). Shaded ribbons represent 95% credible intervals. The Youden-optimal threshold (0.20%) is indicated by a vertical dashed line, corresponding to a sensitivity of 0.767 and specificity of 0.769 (Youden’s J = 0.536; 95% CrI: 0.437-0.624). The adaptive rule-out anchor (T□ = 0.16%; 80% sensitivity target) and rule-in anchor (T□ = 0.48%; 90% specificity target) are indicated by vertical dotted lines, with shading demarcating the three clinical zones: rule-out (dd-cfDNA < 0.16%), indeterminate (0.16%-0.48%), and rule-in (dd-cfDNA > 0.48%). Both anchors fell within the empirically studied range of reported thresholds, confirming that the posterior estimates are clinically grounded. DD-cfDNA, donor-derived cell-free DNA; CrI, credible interval.

**Table 4.** Operating-point ladder for clinical translation. Each row is a candidate dd-cfDNA threshold the analysis supports, with its operating characteristics. The three-zone clinical decision map (Figure 9) uses T1 (rule-out anchor) and T2 (rule-in anchor) as the zone boundaries; Youden sits between them as a balanced reference.

| Operating point | Cut-point (% dd-cfDNA) | Sensitivity (95% CrI) | Specificity (95% CrI) | Clinical role |
| --- | --- | --- | --- | --- |
| **T1 - Rule-out anchor** | 15.57 | 0.80 (target) | 0.742 [0.646-0.826] | Below T1 → rejection unlikely |
| Youden - Balanced | 20.06 | 0.767 | 0.769 | Reference cut-point |
| $\gamma_{\square}$ - Mean obs. thr. | 20.80 (= mean) | 0.764 [0.683-0.831] | 0.775 [0.720-0.822] | Single-threshold summary |
| Marg. summary point | (integrated) | 0.757 [0.678-0.823] | 0.767 [0.713-0.814] | Threshold-marginalised |
| **T2 - Rule-in anchor** | 47.61 | 0.556 [0.429-0.679] | 0.90 (target) | Above T2 → rejection probable; a negative biopsy triggers multimodal work-up, not empiric treatment |

*CrI = 95% credible interval. Cells marked “(target)” are the achieved targets used to define the anchor. The marginalised summary point is integrated over the empirical threshold distribution and does not have a single cut-point. Youden's sens and spec are reported as point estimates because the cut-point itself is the optimisation output; the J statistic at this cut-point carries the posterior uncertainty ( $J = 0.536$ , CrI 0.437-0.624).*

### 6. LOO-CV diagnostics

We assessed out-of-sample predictive performance using Pareto-smoothed importance sampling leave-one-out cross-validation (PSIS-LOO). The blended expected log pointwise predictive density (elpd_loo) was -84.22 (SE = 4.89) and the corresponding

LOO information criterion (LOOIC) was 157.74 (SE = 7.53). Under pure PSIS, the Pareto-k diagnostic exceeded 0.7 for 13 of the 14 studies, which is the expected behaviour in a hierarchical random-effects model with a small number of clusters: each study carries high leverage relative to the parameterisation, and the leave-one-out distributions sit in the tail of the importance-sampling proposal. For those 13 studies, we therefore substituted exact leave-one-out log predictive densities obtained by refitting the model on N - 1 studies and marginalising over the held-out study’s random effects via nested Monte Carlo integration. The blended effective number of parameters was 22.4, reflecting substantial study-level shrinkage.

### 7. Decision curve analysis

We first mapped the net-benefit margin of all five surveillance strategies across the full prevalence (1-50%) and biopsy-threshold space (Figure 6). This revealed that no single strategy is optimal everywhere: biopsy-all is preferred only at high prevalence with a low biopsy threshold, monitor-all only at low prevalence with a high threshold, and the tri-state repeat-if-borderline strategy occupies the clinically central region. The optimal strategy is therefore prevalence- and threshold-dependent rather than fixed.

**Figure 6.**
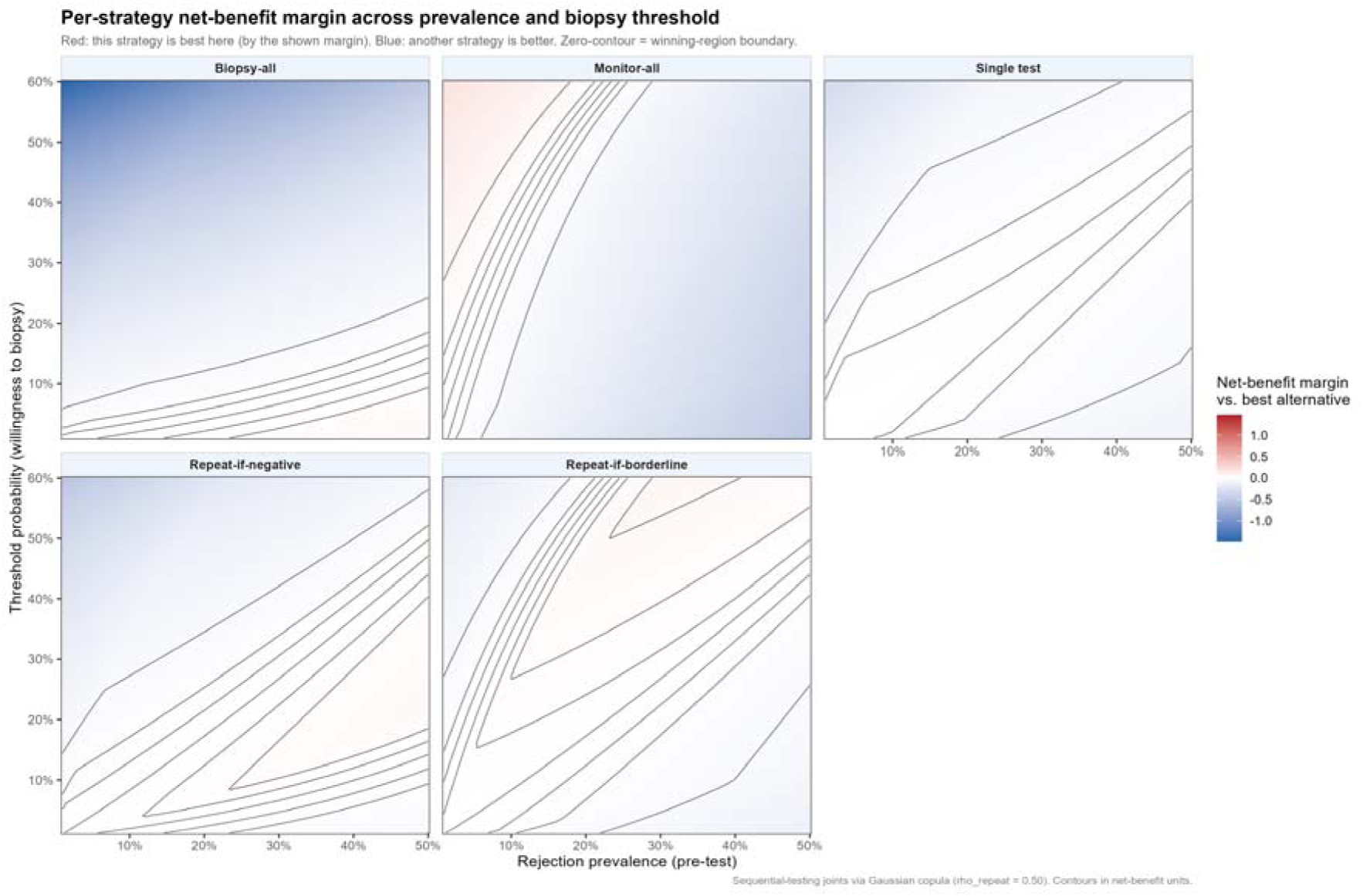
Net-benefit margin for each of five surveillance strategies across rejection prevalence. Net-benefit margin for each of five surveillance strategies across rejection prevalence (x-axis, 1-50%) and biopsy threshold probability (y-axis). Red indicates the strategy is the best available choice; blue indicates another strategy is superior. Biopsy-all is preferred only at high prevalence with a low threshold, monitor-all only at low prevalence with a high threshold, and repeat-if-borderline occupies the clinically central region showing the optimal strategy is prevalence- and threshold-dependent, not fixed. Joint probabilities for sequential-testing strategies were computed using a Gaussian copula with intrapatient correlation ρ = 0.50. Net benefit is defined as sensitivity × π - (1 - specificity) × (1 - π) × [pt/(1 - pt)], where π is prevalence and pt is the threshold probability of biopsy. DD-cfDNA, donor-derived cell-free DNA; CrI, credible interval.

Because the extreme reference strategies (biopsy-all, monitor-all) dominated the color scale in Figure 6 and obscured the differences among the testing strategies, we next restricted the comparison to the three testing strategies alone, each measured against the better of the other two (Figure 7). On this rescaled comparison, the repeat-if-borderline strategy was the best testing option across 62.7% of the plausible prevalence-threshold space, concentrated at low-to-intermediate prevalence, whereas a single test at the Youden cutoff was never optimal at any point. Where the strategies were near-indistinguishable (near-white regions), the choice among them was not clinically consequential.

**Figure 7.**
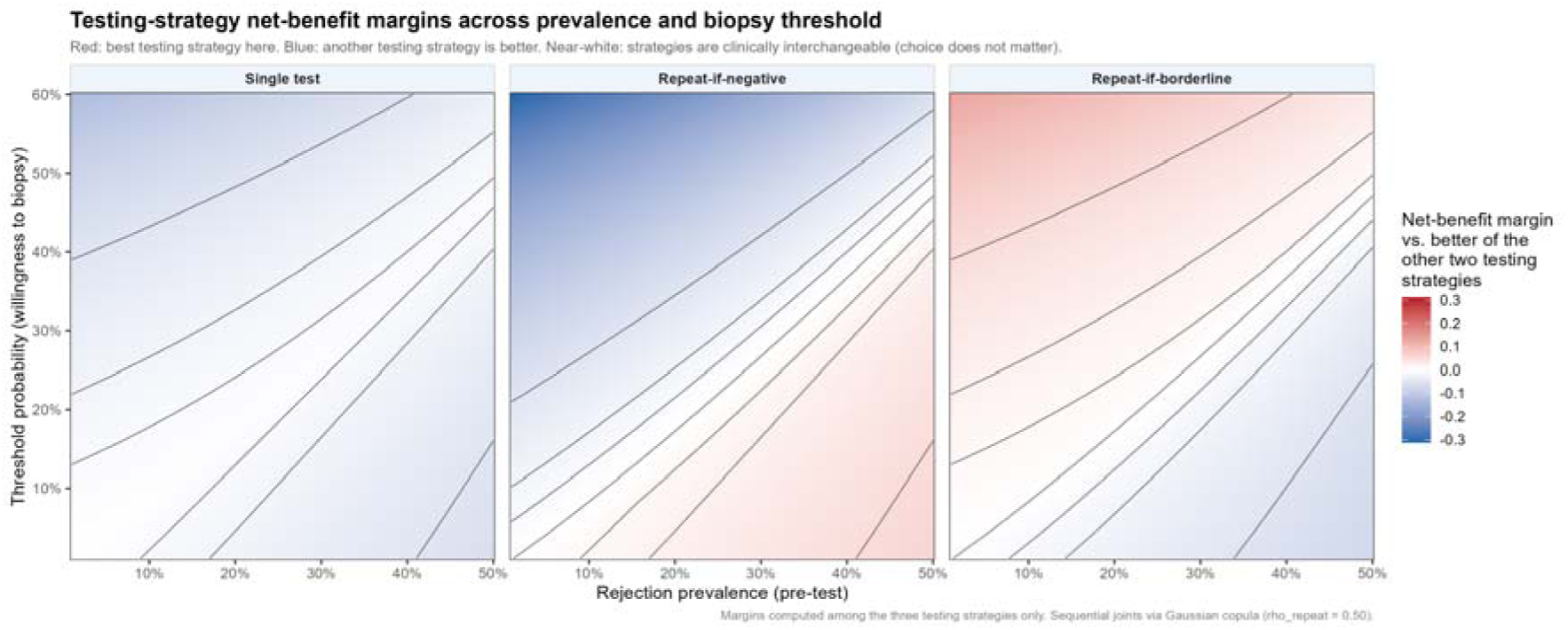
Testing - Strategy net benefit margins across prevalence and biopsy threshold. Net-benefit margin among the three testing strategies (each versus the better of the other two). Near-white regions indicate the strategies are practically interchangeable; saturated regions indicate a material advantage. Repeat-if-borderline is the best testing option across 62.7% of the plausible prevalence-threshold space, concentrated at low-to-intermediate prevalence.

Finally, because a relative margin shows which strategy is best but not whether it is worth doing, we examined each testing strategy’s absolute net benefit (Figure 8). Repeat-if-borderline maintained positive net benefit across 82.1% of the space, the widest safe operating range of any strategy evaluated, and was net-harmful only in the low-prevalence, high-threshold corner, precisely the asymptomatic surveillance setting in which no testing-and-acting strategy adds value and a high-negative-predictive-value rule-out approach is appropriate. Its advantage was concentrated at low-to-intermediate prevalence and narrowed at high prevalence, where a first elevated result is usually true and the strategies converge.

**Figure 8.**
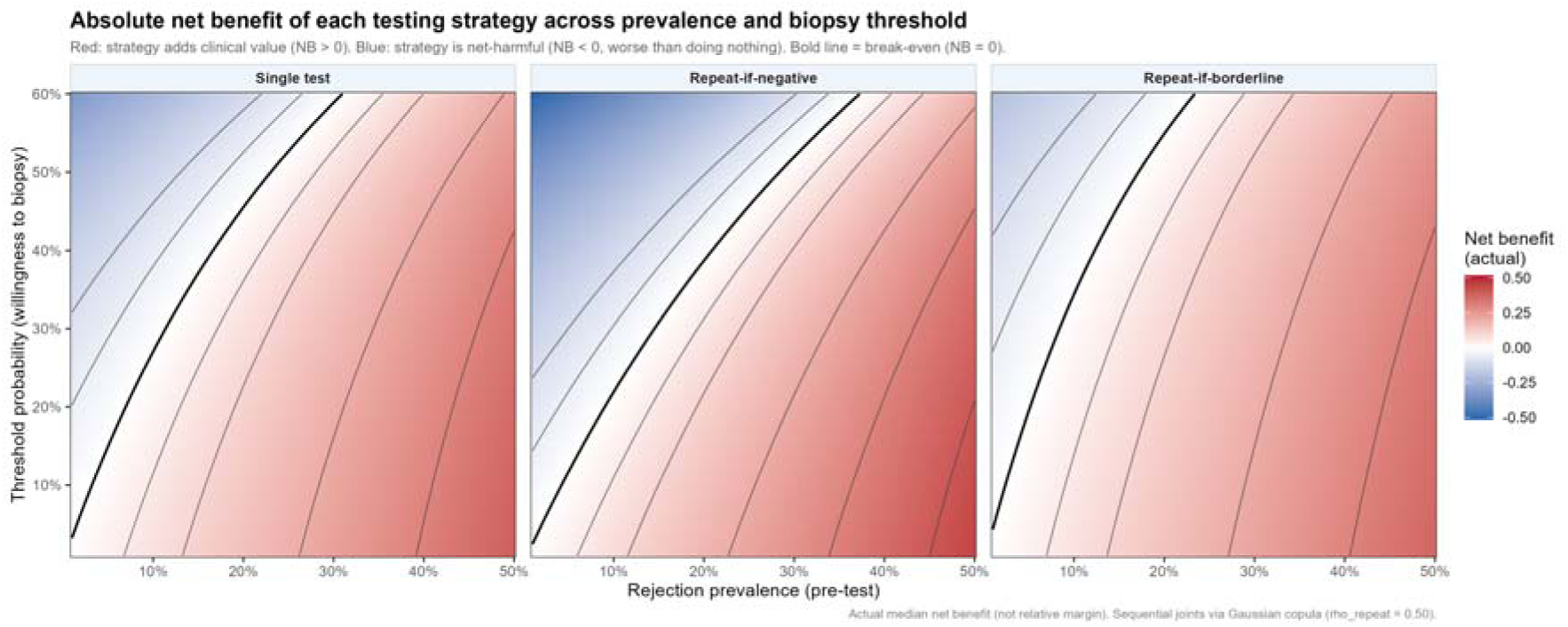
Absolute net benefit for each testing strategy across prevalence and biopsy threshold. Absolute net benefit of each testing strategy across rejection prevalence and biopsy threshold. Red indicates positive net benefit (adds clinical value); blue indicates net-harmful (net benefit < 0); the bold line marks break-even. All three testing strategies are net-harmful only in the low-prevalence / high-threshold corner, the asymptomatic surveillance setting, where a high-negative-predictive-value rule-out approach is appropriate. Repeat-if-borderline maintains positive net benefit across 82.1% of the plausible space, the widest safe operating range of any strategy. Sequential-testing joints via Gaussian copula (ρrepeat = 0.50).

### 8. DCA robustness to within-patient correlation

The net-benefit ranking and the advantage of the tri-state repeat-if-borderline strategy were preserved across sequential-testing correlations *ρ* ∈ {0.30,0.50,0.70} the strategy remained the best testing option across 61.0-64.0% of the prevalence-threshold space and net-beneficial across 81.0-83.2%, with the mean net-benefit margin varying by less than 0.005 units. The decision-curve conclusions are therefore insensitive to the assumed within-patient correlation.

### 9. Robustness to clustering assumption

To assess robustness, we re-fitted the full HSROC model under ρ ∈ {0.0, 0.3, 0.5, 0.7}. All primary estimates were highly robust across the ρ range.

- AUC varied between 0.779 and 0.784 (range = 0.005)
- Youden cutoff varied between 0.194% and 0.204% (range = 0.010 percentage points)
- 80% sensitivity anchor cutoff varied between 0.146% and 0.159%
- 90% specificity anchor cutoff varied between 0.476% and 0.489%
- Marginalised summary sensitivity varied between 0.753 and 0.759; specificity between 0.765 and 0.772

### 10. Heterogeneity, Publication Bias, and Age Meta-Regression

The between-study variance in diagnostic accuracy 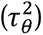 was 0.306 (95% CrI: 0.109-0.969) and the variance in the threshold effect 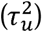 was 0.493 (95% CrI: 0.140-1.801).

Deeks’ funnel plot asymmetry test yielded a slope of +2.90 (p = 0.0106) for the regression of the log diagnostic odds ratio on the inverse root of the effective sample size.

A continuous Bayesian meta-regression of the HSROC location parameter on study-level centred median age produced a coefficient (*β_age_*) of -0.0158 (95% CrI: -0.0512 to 0.0159).

## Discussion

In this analysis, we sought to provide a clinically grounded decision framework for post-transplant surveillance and determine whether a single threshold could be clinically meaningful.

Youden’s index, which we considered as the baseline, due to its mathematical assumption of equal sensitivity and specificity, this optimal threshold was identified at 0.20% with a sensitivity of 0.77 and specificity of 0.77; this implies that a randomly sampled rejection episode had approximately a 78% probability of yielding a higher dd-cfDNA value than a randomly sampled non-rejection episode; however, almost a quarter of rejections could not be caught. This operating point is therefore a valid summary of discrimination rather than a directly deployable decision rule, since screening demands high sensitivity and confirmation demands high specificity. Therefore, we conclude that a single threshold cannot serve both roles simultaneously.

This observation should be interpreted strictly within the framework set out in the Methods. The 0.20% Youden threshold is not an incorrect answer to the question it was constructed to address. As a classification benchmark it correctly summarises the intrinsic discriminatory capacity of the biomarker, and its derivation neither assumed nor required a prevalence. What our analysis demonstrates is that this quantity does not transport to the second question. A cutoff selected to maximise the sum of sensitivity and specificity, weighting a missed rejection episode and an unnecessary biopsy equally, has no mathematical reason to coincide with the cutoff that maximises net benefit in a surveillance population in which rejection prevalence may be below 2% and in which the two errors carry markedly unequal consequences. The divergence we observe between the Youden-optimal point and the decision-analytically preferred strategies is therefore the expected consequence of changing the question, and not evidence that the earlier threshold was derived in error. We emphasise this distinction because the existing literature has generally been rigorous with respect to classification and comparatively silent with respect to deployment; the gap our framework addresses lies in the latter.

The adoption of single-threshold approach is underscored by inherent structural biases in literature. Our detection of significant small-study effects via Deeks’ asymmetry test (p = 0.0106) exposes that early, localized pilot studies have systematically reported overly optimistic diagnostic accuracies. This artefactual inflation is a known consequence of overfitting rigid binary thresholds to underpowered cohorts to maximize early-phase diagnostic odds ratios. When these highly optimized cutoffs are extrapolated to broader, heterogeneous populations, they become predictively volatile, this highlights the clinical danger of anchoring any surveillance protocols to a single threshold approach. Furthermore, the substantial threshold variance [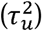 = 0.493] identified in our primary HSROC architecture mathematically demonstrates that relying on a static diagnostic boundary across differing clinical environments is inherently flawed. Crucially, our continuous Bayesian meta-regression confirmed that this variance is not merely an artifact of pooling pediatric and adult cohorts. The failure of the age covariate to stratify the threshold variance [(*β_age_*) CrI: - 0.0512 to 0.0159)] reflects the complex, blended reality of modern transplant centers, wherein pediatric programs routinely manage adult-sized Adult Congenital Heart Disease (ACHD) patients, and adult programs evaluate early adolescents. Because baseline shedding kinetics and absolute blood volumes are inextricably blurred across these developmental stages at the study level, the pursuit of a singular, demographic-specific binary threshold is clinically unhelpful.

A necessary corollary of this argument is that the between-study variance we report must be attributable to something, and the most immediate candidate warrants direct examination: that different commercial platforms simply measure different things, such that pooling them manufactures heterogeneity as an artefact. The available head-to-head evidence does not support this interpretation, and the point is material because the reading of our entire model depends upon it.

Two studies have compared modern single-nucleotide polymorphism (SNP) based platforms on identical specimens. Rodgers et al. analysed 428 paired samples from 112 heart transplant recipients using standard and expanded SNP panels and found no significant difference between them in the detection of acute rejection, alongside improved specificity relative to gene expression profiling at unchanged sensitivity (36). Hsi et al. performed the corresponding comparison between AlloSure (CareDx) and Prospera (Natera) across 248 consecutive, simultaneously drawn samples; both assays were correctly suggestive of rejection in every instance in which a contemporaneous biopsy was available, and the discordance that did occur was confined to 23 of 248 samples and was attributable to the differing manufacturer-recommended reporting thresholds, 0.12% and 0.15% respectively, rather than to any divergence in the underlying molecular quantification (37). Taken together, these data indicate that the analytical performance of contemporary SNP-based dd-cfDNA assays is closely comparable, and that where the platforms disagree they do so at the level of the interpretive threshold applied to the result rather than at the level of the measurement itself.

This carries a direct consequence for the interpretation of our model. If the assays measure the same underlying biology to a comparable analytical standard, then the substantial between-study variance in our HSROC architecture, comprising a threshold-effect variance of 0.493 (95% CrI 0.140 to 1.801) and a location variance of 0.306 (95% CrI 0.109 to 0.969), cannot be dismissed as measurement artefact arising from the pooling of incommensurable platforms. Neither, as demonstrated above, is it explained by the age composition of the constituent cohorts. What remains is variation in how the test is deployed: the post-transplant interval at which surveillance samples are drawn, the sampling cadence, whether the paired biopsies are protocol-driven or for-cause, the immunosuppression regimen and the point in the taper at which testing occurs, the institutional threshold for referral to biopsy, and the baseline rejection risk of the population served. These are properties of clinical environments, not of reagents.

This inference converts our central recommendation from a statistical preference into a structural necessity. Were the heterogeneity to originate in assay mechanics, the appropriate remedy would be technical: harmonise the platforms, establish a universal calibrator, and a single transportable threshold would follow in due course. Because it originates instead in clinical implementation, no degree of analytical harmonisation can yield a universal threshold, since the quantity that varies is not what the assay measures but the population and protocol to which the measurement is applied. Local calibration is therefore not a concession to imperfect evidence but the logically required response to the structure of the variance we observe, and it is on this basis that we advance the tri-state architecture as a blueprint to be instantiated locally rather than as a set of numerical anchors to be adopted directly.

Ultimately, dd-cf-DNA cannot replace an endomyocardial biopsy, detecting the specific pathophysiology of allograft injury is beyond the biomarker’s biological capacity. This is particularly relevant given the vastly divergent shedding kinetics of different rejection phenotypes. In the GRAfT study (24), antibody-mediated rejection (AMR) produced higher and more sustained dd-cfDNA elevations (median ∼1.68% for grade ≥2), with a longer lead time and detectable elevation preceding diagnosis in the majority of cases (12/15), reflecting diffuse microvascular injury; acute cellular rejection (ACR) produced lower-magnitude, briefer elevations (median ∼0.33% for grade ≥2) that preceded diagnosis in only a minority of cases (2/17). AMR is not uniformly catastrophic but follows a variable course; it may be subclinical or clinically silent yet still adversely affect prognosis (28) Consequently, a static threshold such as 0.20% is oversensitive for severe AMR yet inadequately calibrated for early ACR, and the biomarker’s highest utility is fundamentally that of a triage tool.

The necessity of transitioning from a binary threshold to a tri-state triage framework is not without precedent in cardiovascular medicine. The evolution of dd-cfDNA interpretation closely mirrors the trajectory of high-sensitivity cardiac troponin (hs-cTn). Just as the cardiology community recognized that forcing a continuous marker of myocardial injury into a single binary cutoff generated an epidemic of false positives, prompting the European Society of Cardiology to formally adopt the 0h/1h Rule-Out/Rule-In algorithm, we recommend that the thoracic transplantation must similarly adapt to its high-sensitivity molecular assays.(27)

Real-world institutional practices have already begun to bypass the standard threshold in recognition of these limitations. In the broader solid-organ transplantation, the use of dd-cf-dna in recent large-scale observational cohorts are have actively evaluated elevated surveillance thresholds up to 1.0% to accurately capture rejection severity, (29) while multimodal diagnostic frameworks like the Two-Threshold Algorithm (2TA) have successfully integrated absolute quantification (copies/mL) alongside fractional percentages to drastically reduce false-positive rates. (30) Our derived tri-state anchors provide the formal biostatistical validation for this ongoing clinical evolution.

The convergence of substantial heterogeneity, small-study publication bias, and the clinical reality of ACHD demographics collectively argue against the utility of the single-threshold paradigm. These datapoints support transition toward the adoption of our proposed tri-state framework, serving as a validated architectural blueprint that empowers institutions to locally calibrate a highly sensitive Rule-Out anchor, protecting healthy, asymptomatic patients from iatrogenic harm, and a highly specific Rule-In anchor to triage high-risk patients for definitive biopsy.

Our adaptive walk-down procedure, after pooling all available evidence, reported the highest sensitivity and specificity the data could realistically support, showcased a rule-out threshold of 0.16% at 80% sensitivity and a rule-in threshold of 0.48% at 90% specificity. Our framework identifies three clinically actionable zones (Figure 9). The posterior indicates that approximately one in five rejection episodes remains undetectable even at an aggressively low 0.16% cutoff, a ceiling likely driven by non-shedding rejection phenotypes and assay noise floors, rather than by any limitation of the statistical model. Consequently, dd-cfDNA cannot be relied upon as a standalone screening or confirmatory tool, and the 0.32-percentage-point indeterminate zone between the anchors is not a failure of the analysis but a direct reflection of the biomarker’s biological limits. Results falling in this zone require contextualisation with secondary signals such as echocardiography, gene expression profiling, or donor-specific antibody testing.

**Figure 9.**
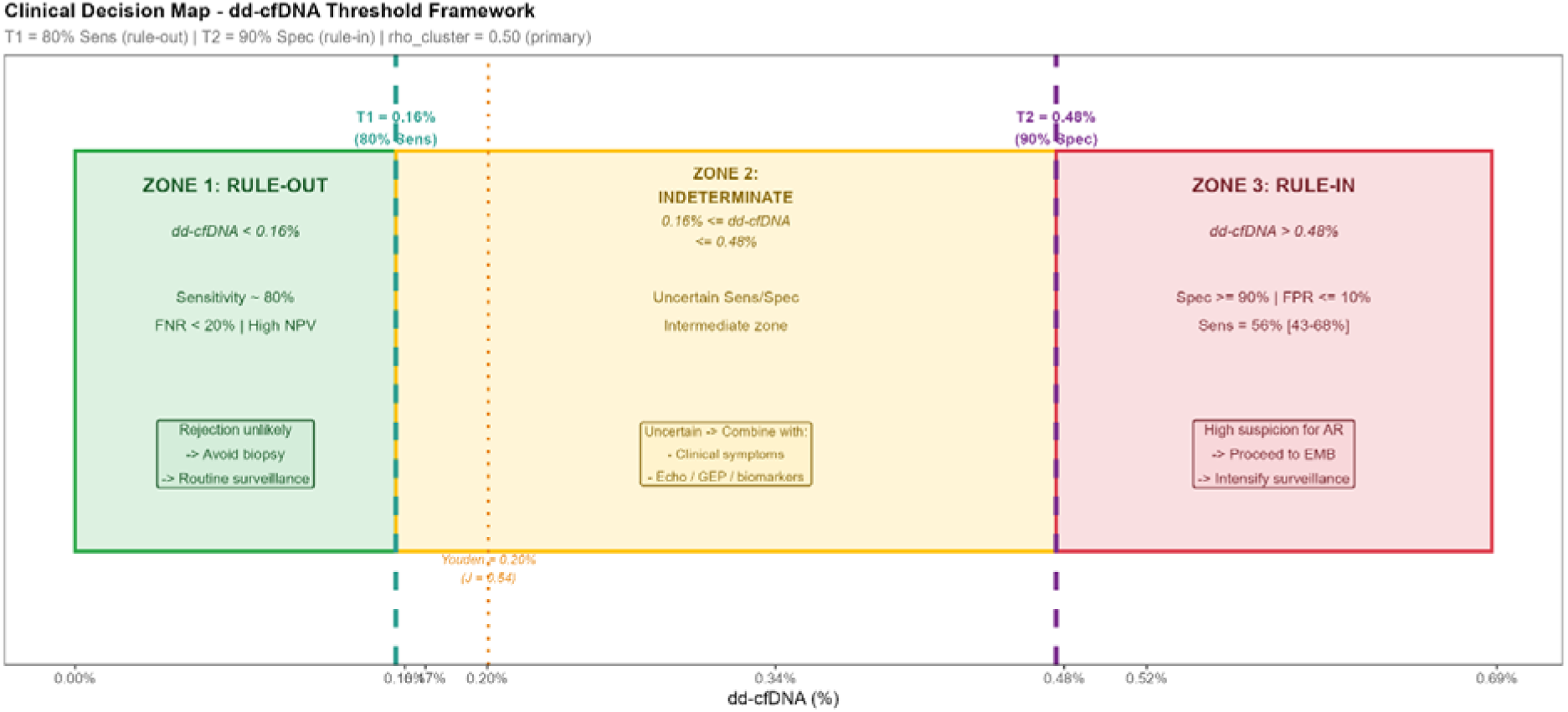
Proposed tri-state clinical decision framework for dd-cfDNA-guided post-transplant surveillance. Three clinically actionable zones are defined by the adaptive rule-out threshold (T□ = 0.16%; 80% sensitivity anchor) and the rule-in threshold (T□ = 0.48%; 90% specificity anchor). Zone 1 (dd-cfDNA < 0.16%): rejection unlikely; routine surveillance appropriate. Zone 2 (0.16%-0.48%): indeterminate/grey zone; clinical synthesis with secondary testing (echocardiography, gene expression profiling, donor-specific antibodies) or repeat testing within 3-7 days is recommended. Zone 3 (dd-cfDNA > 0.48%): rejection probable; strong consideration of endomyocardial biopsy and multimodal investigation; a discordant negative biopsy prompts deeper histological work-up, donor-specific antibody testing, functional imaging and repeat dd-cfDNA within 1-2 weeks rather than empiric treatment, since treatment decisions rest on tissue diagnosis and the integrated clinical picture rather than on the biomarker alone. The Youden-optimal threshold (0.20%) is shown for reference. DD-cfDNA, donor-derived cell-free DNA; EMB, endomyocardial biopsy.

### Why one in five rejection episodes falls below the rule-out anchor

The residual false-negative rate of approximately 20% applies directly at the rule-out anchor, T = 0.16%, the cut-off at which the pooled posterior sustains 80% sensitivity. The adaptive walk-down tested descending sensitivity targets beginning at 95%; 80% was the highest target that at least half of the posterior draws could sustain across the 14 included cohorts (1,763 patients). Modelled sensitivity continues to rise at cut-offs below 0.16%, but the accompanying loss of specificity makes lower cut-offs clinically impractical as rule-out tools, and because no included study applied a threshold below 0.10%, cut-points below that value lack empirical support. The 80% figure is therefore the sensitivity ceiling that the published evidence can realistically support within the studied range, not an arbitrary biological constant. Four mechanisms, acting in combination, account for the rejection episodes that fall below it.

The first is the biology of donor DNA release. dd-cfDNA is the end product of allograft parenchymal cell death by necrosis and apoptosis, which releases 120-160 bp nucleosomal fragments into the circulation. In cellular rejection the infiltrating mononuclear cells are recipient-derived and release no donor DNA. Under the ISHLT 2004 working formulation, grade 1R is defined as interstitial or perivascular infiltration with at most one focus of myocyte damage (38); where parenchymal necrosis is minimal or absent, donor DNA release remains correspondingly low. The prospective cohorts confirm this biological gradient. In GRAfT, median dd-cfDNA was 0.02% in non-rejection (grade 0) and 0.04% in ACR 1R, barely above baseline, rising nine-fold to 0.34% in ACR ≥2R (24). In the DEDUCE cohort, the medians for non-rejection (0.04%) and ACR 1R (0.045%) were practically indistinguishable, whereas pAMR1(I+) yielded 0.44% and pAMR2 3.43% (13). Low-grade cellular infiltration frequently occurs without appreciable parenchymal death, and the assay cannot detect what the graft does not shed.

The second concerns the clinical significance of the episodes that are missed. Under the ISHLT guidelines for the care of heart transplant recipients, grade 0R and grade 1R cellular rejection are managed expectantly without augmentation of immunosuppression (39). To the extent that non-shedding episodes are enriched for low-grade, non-cytolytic events, the sensitivity of dd-cfDNA for clinically actionable rejection is higher than the composite 80% implies. That reassurance should not be over-read, however, because actionability is not an isolated histological property. Kim et al. documented rare ACR 2R episodes with preserved left ventricular ejection fraction (≥60%) that shed silently (≤0.02%) across serial measurements (13); conversely, an asymptomatic grade 1R episode in a sensitised recipient with new de novo donor-specific antibodies or declining left ventricular global longitudinal strain may warrant clinical intervention. Because actionability emerges from the complete clinical picture, dd-cfDNA cannot serve as a standalone arbiter of treatment; its role is that of an initial triage filter.

The third is that a proportion of these apparent misses are errors of the reference standard rather than of the biomarker. Endomyocardial biopsy is an imperfect gold standard, and pathologist agreement on biopsy grading is limited. In CARGO II, for an average pair of pathologists, fewer than one-third of biopsies read as ≥2R by one reader were confirmed by the other, and all-grade concordance of approximately 71% was driven almost entirely by grade 0 (40); a formal reproducibility study using the 2004 criteria reported a combined Cohen’s kappa of only 0.39 (41). Benign morphological mimics compound this.

Subendocardial Quilty B lesions were present in 35% of centrally downgraded biopsies and triggered inaccurate rejection diagnoses in 3-5% of all CARGO biopsies (42), and reparative granulation tissue at prior bioptome sites is likewise over-read on light microscopy as grade 1R or 2R. In these instances the biopsy is a false positive and the uninjured allograft appropriately sheds no dd-cfDNA; our model, obliged to treat histology as infallible, records the biomarker as having missed a rejection that did not occur.

The fourth is sampling error, which operates in the opposite direction for focal disease. In post-mortem examination of hearts with patchy lymphocytic myocardial infiltration, obtaining even 10 biopsy samples per ventricle yielded a false-negative rate of 37-45% (43); the reference standard is therefore both less sensitive and less specific than a composite endpoint assumes. Finally, and as developed at length below, because commercial assays report a fraction, donor/(donor + recipient), intercurrent infection, surgical wound healing or systemic inflammatory stress elevates total recipient cfDNA and mechanically suppresses the donor fraction below 0.16% despite active allograft injury (30). Taken together, these mechanisms locate the residual false-negative rate in the biology of shedding and in the limitations of histology rather than in the statistical model, and they underline that a result below TL lowers, but does not abolish, the probability of rejection.

To evaluate whether these thresholds translate into actionable clinical benefits, we employed decision curve analysis (DCA), which grounds biomarker performance in clinical reality by weighing the harm of an unnecessary biopsy against the benefit of detecting rejection. Across the full modelled prevalence range (1-50%), dd-cfDNA-guided strategies demonstrated positive net benefit over both the biopsy-all and monitor-all reference strategies except in the low-prevalence, high-threshold region where no testing strategy adds value.

To ground these thresholds in clinical reality, we evaluated net benefit by decision-curve analysis across the full clinically relevant range of rejection prevalence (1-50%), from surveillance to for-cause and DSA-positive antibody-mediated rejection populations. This range is supported by the included evidence itself: per-test rejection prevalence, recomputed from the study-level 2×2 data on a consistent per-sample basis, ranges from approximately 0.8% to 52% across the 14 studies. The large multicenter cohorts cluster at the low end (∼0.8-7%; (13, 16, 19,24, 26), consistent with contemporary surveillance series (∼1.2% treated acute rejection) (35), while the high-prevalence tail comprises smaller single-center and pediatric cohorts. Per-test prevalence in the largest cohorts therefore sits well below the pretest probabilities typically invoked to characterize the biomarker, with direct implications for its surveillance use.

Taken together, these analyses show that the tri-state repeat-if-borderline strategy is both the most frequently optimal testing strategy (best across 62.7% of the space) and the safest (net-beneficial across 82.1%, the widest safe operating range of any strategy). Its advantage is concentrated at low-to-intermediate prevalence and narrows at high prevalence, where a first elevated result is usually true and the strategies converge. Critically, the only region in which no testing strategy adds value is the low-prevalence / high-threshold corner, which corresponds precisely to the asymptomatic surveillance setting and reinforces that the biomarker’s role there is rule-out rather than rule-in. At high prevalence, where a first elevated result is usually true, the marginal value of retesting an indeterminate result before acting is low, and the temporal risk of delaying intervention in a rapidly failing graft, which the net-benefit calculation does not itself capture, may favor proceeding directly to biopsy (Figures 6-8).

Ultimately, our DCA modelling proves that the optimal use of dd-cfDNA cannot be rigidly fixed to a single mathematical point. The traditional paradigm of seeking one perfect threshold, such as the Youden index, is fundamentally flawed by the continuous nature of the biomarker. When forced into a binary switch, the nuance is not fully captured. The biological reality strongly favors a continuous, dynamic interpretation, however, the historical persistence of the single-threshold paradigm must be understood as a necessary byproduct of early clinical translation. To establish initial clinical utility and align with the dichotomous diagnostic frameworks traditionally required for regulatory evaluation, it was pragmatically necessary to operationalize a continuous biomarker into a definitive binary switch. While essential for early implementation, our DCA showcases that persisting with this rigid dichotomization strips away critical biological nuance. Our findings strongly advocate for a structural transition away from rigidly applied single-threshold framework. Instead, we propose the adoption of a tri-state, prevalence-aware framework. The 0.32 percentage point grey zone between 0.16% and 0.48% is biological reality, and clinicians should dynamically toggle between maximizing specificity to protect healthy patients and maximizing sensitivity to rescue failing allografts. This gray zone is not a diagnostic failure; it is active space meant for clinical rationale and decision making synthesizing secondary signals, such as gene expression profiling (GEP), echocardiography, and the trajectory of serial testing.

The biomarker performance depends entirely on the chosen threshold, and no single point represents the entire curve. The close agreement between the marginalized summary and the γL operating point indicates that the empirical threshold distribution is approximately symmetric around its mean. The wide credible interval on AUC (lower bound 0.668) reflects substantial between-study heterogeneity, which the HSROC framework explicitly accommodates by modelling threshold variation as a covariate rather than treating it as noise.

Our work has limitations; there are differences in the various platforms (AlloSure, in-house PCR, Shotgun PCR), and we did attempt to formally incorporate a covariant modelling; however, there was overfitting due to the small number of studies, and it would be advisable to prospectively validate it with specific assay types. As our work is a meta-analysis, our thresholds are population-level and should be calibrated locally. Importantly, while our framework identifies specific rule-out (0.16%) and rule-in (0.48%) anchors, these absolute values are not golden bullets. The data explicitly demonstrates that clear, universal binary decisions do not exist in the context of this continuous biomarker. Because there is currently no universal calibrator across differing technologies, pooling data from disparate assays inherently aggregates distinct biological baselines. Consequently, our derived thresholds must be viewed as conceptual placeholders that validate the tri-state architecture. For real-world implementation, individual transplant centers must utilize this architectural blueprint to locally calibrate their specific assays, partnering with institutional statisticians to identify their own localized high-NPV and high-PPV anchors. We applied a design effect correction, and the effective sample size was reduced to 2,518 tests, which was conservative; however, it helped retain the robustness of the study. The retest time was assumed to be 3-7 days, which is clinically informed; however, it was not empirically validated for the copula model.

Fractional confounding of dd-cfDNA is a further limitation, and a central concern of contemporary practice, the dd-cfDNA fraction reflects both donor-derived and recipient-derived cfDNA. Absolute donor quantification captures graft injury more directly than the fraction alone, which is the rationale for combined fraction-plus-quantity approaches such as the two-threshold algorithm of Kim et al. (30). The pooled thresholds analyzed here are fractional and therefore subject to this confound. We emphasize, however, that our contribution is a decision architecture : rule-out, indeterminate, and rule-in zones with selective repeat testing, rather than a specific universal cutoff; this architecture is agnostic to the underlying quantification method and can be recalibrated as absolute-quantity data accumulate.

Two considerations deserve fuller treatment here, because they pull in opposite directions and the balance between them is not yet settled in the field.

The first is that the fractional metric possesses genuine and frequently underappreciated pre-analytical advantages. Expressing donor DNA as a proportion of total cell-free DNA constructs a self-normalising ratio in which the donor and recipient compartments share a common denominator and are consequently subject to identical ex vivo handling. Leukocyte lysis within the blood collection tube, which releases recipient genomic DNA into the plasma and constitutes the dominant pre-analytical artefact in cell-free DNA measurement, inflates numerator and denominator within the same specimen and therefore largely cancels in the ratio. The same holds for variability in extraction efficiency, for differences in plasma volume and centrifugation protocol, for delays between venepuncture and processing, and for library preparation yield. An absolute measurement expressed in copies per millilitre enjoys none of this protection: each of these variables acts directly and unopposed upon the reported figure, and absolute quantification consequently imposes considerably stricter demands on specimen handling, standardised collection tubes, and inter-laboratory calibration. This robustness to technical sources of variation is a principal reason the fraction was adopted for initial clinical validation, and we do not dispute its value on these grounds.

The countervailing consideration is that the same denominator conferring pre-analytical stability simultaneously introduces biological instability. The total cell-free DNA background of the recipient is not a constant; it is a dynamic reflection of systemic leukocyte turnover.

Neutrophils and lymphocytes are short-lived and are the principal contributors to circulating cell-free DNA in health, and any process accelerating their turnover raises the background accordingly. Localised infection, uncomplicated post-operative recovery, physical trauma, strenuous exertion, and ordinary physiological stress each measurably elevate total cell-free DNA without any change whatsoever in the donor organ. Because the donor signal occupies the numerator, an elevated recipient background mathematically suppresses the reported fraction. The consequence is directional and clinically adverse: the suppression occurs precisely in those patients who are systemically unwell, which is the population in whom rejection is most likely to be under active consideration. A recipient with concurrent pneumonia and active cellular rejection may therefore return a fraction below the rule-out anchor while shedding an unambiguously abnormal quantity of donor DNA. This is a false negative generated by arithmetic rather than by any failure of the assay to detect donor molecules.

We accept the argument that molecular surveillance assays were validated in, and are intended for, clinically stable recipients, and that testing should ordinarily be deferred in the setting of sepsis or major haemorrhage. Our concern lies with the intermediate range. Sepsis and haemorrhage are conspicuous and prompt deferral; a resolving urinary tract infection, a recent line insertion, or a modest inflammatory response is not conspicuous, is not ordinarily regarded as grounds for deferral, and may nonetheless perturb the denominator sufficiently to displace a result across an anchor. Denominator confounding is thus better characterised as a continuously distributed bias than as a rare edge case. It is furthermore unobservable from the reported fraction alone, since a single fractional value cannot distinguish a genuinely low donor signal from an elevated recipient background.

Absolute quantification addresses this directly by removing the recipient from the measurement. Reporting donor genome equivalents per millilitre isolates the quantity of clinical interest, namely the rate at which the allograft is releasing DNA, from the systemic state of the recipient, at the acknowledged cost of forfeiting the pre-analytical cancellation described above. Neither metric therefore dominates the other, and the appropriate resolution is not to select between them but to report both. The two-threshold algorithm of Kim et al. (30) provides the structural template: a fractional threshold and an absolute quantity threshold are applied in parallel, and a result exceeding either is treated as abnormal. Discordance between the two carries interpretable information in its own right, since a low fraction accompanied by a high absolute quantity is the specific signature of denominator inflation. We regard this parallel architecture, rather than the wholesale replacement of one metric by the other, as the probable structural future of the field. Our tri-state framework is compatible with it: the rule-out, indeterminate, and rule-in architecture is defined by the clinical purpose of each zone rather than by the units in which the biomarker is expressed, and the same three zones may be instantiated on an absolute or a dual-metric scale as calibration data accumulate.

Change in dd-cfDNA fraction over time. Because the measurement is fractional, non-rejection changes in the recipient cfDNA background can shift the ratio independently of donor shedding. This is particularly relevant in the first post-transplant year, when immunosuppression is actively adjusted, prednisone tapering and reductions in tacrolimus targets alter recipient leukocyte turnover and cfDNA background, and can raise or lower the dd-cfDNA fraction with no change whatsoever in donor-derived shedding. This temporal confounding further motivates absolute donor quantification and trajectory-based (serial-delta) interpretation rather than reliance on any single fractional time-point.

The persistently indeterminate result. A common and under-addressed scenario, directly analogous to high-sensitivity cardiac troponin, is the repeat test that again falls between the rule-out and rule-in anchors. Our framework does not by itself resolve this case. We regard the persistently indeterminate result as an explicit indication for clinical synthesis rather than reflexive biopsy, and as a priority for future work aimed at reducing residual ambiguity including serial-delta (trajectory) interpretation and integration with secondary signals such as gene-expression profiling, echocardiography, and donor-specific-antibody testing.

### The biomarker-positive, tissue-negative result

dd-cfDNA was introduced principally as a rule-out test that permits a protocol biopsy to be skipped, the purpose of a rule-in anchor deserves explicit statement, and the clearest way to give it is through the case that most often arises in practice. Consider an asymptomatic, haemodynamically stable recipient with a dd-cfDNA of 0.50% whose endomyocardial biopsy shows neither acute cellular nor antibody-mediated rejection (ISHLT grade 0R / pAMR0). This patient should not receive empiric anti-rejection therapy. We do not recommend initiating pulsed corticosteroids or cytolytic agents on the strength of an isolated molecular biomarker against negative histology (39); doing so would expose the patient to serious iatrogenic toxicities, including opportunistic infection, CMV pneumonitis, nephrotoxicity and post-transplant lymphoproliferative disorder, without a confirmed target pathology.

Bayesian decision theory supports this conservative stance. At the rule-in anchor (TL = 0.48%; specificity 0.90, sensitivity 0.556; pooled positive likelihood ratio ≈ 5.6), the positive predictive value is approximately 38% at a surveillance prevalence of 10% and approximately 70% at a prevalence of 30%. Because the majority of results above TL at surveillance prevalence are not acute rejection, “rule-in” in our Bayesian framework signifies a requirement for invasive tissue diagnosis and multimodal investigation, not a mandate for treatment. Its value lies in the direction of the error it prevents: a result above TL should never be dismissed on the strength of a single negative biopsy. Our analytical architecture is obliged to score such events as false positives, because a diagnostic test accuracy meta-analysis treats histology as an infallible reference standard. That mathematical necessity should not be mistaken for a biological conclusion. Several mechanisms make it likely that a substantial proportion of these results reflect genuine allograft injury which the reference standard has failed to capture.

The first is sampling. A standard endomyocardial biopsy procures only three to five fragments of 1-2 mm from the right ventricular septum, a minute and non-random sample of a ventricle exceeding 100 g, whereas dd-cfDNA integrates DNA release across the entire graft. Cellular rejection is frequently patchy, and focal infiltrates situated away from the sampled septal region are simply not present within the specimen retrieved; in the post-mortem series cited above, even 10 samples per ventricle missed patchy lymphocytic infiltration in 37-45% of cases (43). The reported interobserver variability in the histological grading of rejection compounds this, since the same tissue may be graded differently by different pathologists (14, 40, 41). A negative biopsy is therefore evidence of absent rejection at the sites sampled, which is a materially weaker statement than absent rejection within the graft.

The second is temporal. Molecular injury precedes histological change. Donor DNA is released when donor cells die, and detectable release may therefore begin before the cellular infiltrate or the microvascular changes required to satisfy histological diagnostic criteria have accumulated. The GRAfT data are consistent with this, with elevation preceding formal diagnosis in the majority of antibody-mediated rejection episodes (24). A biopsy performed at the moment of an elevated result may thus be sampling a process that has commenced molecularly but has not yet become histologically diagnosable, and a result classified as a false positive against the concurrent biopsy may prove to have been an early true positive against a biopsy performed one to two weeks later.

The third is that dd-cfDNA is a marker of allograft injury rather than of rejection specifically. Injury arising from causes other than alloimmune attack, including ischaemia-reperfusion in the early post-operative period, cardiac allograft vasculopathy, infection involving the graft, and recent instrumentation including the biopsy procedure itself, releases donor DNA without producing rejection histology. These are true positives for graft injury and false positives only against the narrower endpoint by which our model scores them. Antibody-mediated activity that is serologically and molecularly evident before it becomes histologically manifest belongs to the same category, and biopsy-negative antibody-mediated rejection is now a recognised clinical entity.

Recent prospective data indicate that an elevated dd-cfDNA with a negative biopsy is rarely benign noise. In the GRAfT cohort, among 46 heart transplant recipients with elevated dd-cfDNA (median 0.49%, virtually identical to our rule-in anchor) and biopsies read as negative for rejection, 59% developed subsequent acute rejection, allograft dysfunction or death within one year, compared with 20% of patients with a positive biopsy but a negative dd-cfDNA (14). Correspondingly, the positive likelihood ratio of dd-cfDNA in GRAfT was 2.59-3.74 when judged against isolated histology alone, a figure depressed by local-versus-core pathologist discordance and lower than our pooled aggregate posterior estimate of approximately 5.6 across 14 cohorts, but rose to 7.63-9.82 when evaluated against the composite one-year clinical outcome (14). The “false positive” against the biopsy was, in the majority of cases, a true positive against the patient.

When the dd-cfDNA is 0.50% and the biopsy is grade 0, therefore, the rule-in anchor exists to prevent false reassurance. The clinician neither treats empirically nor dismisses the finding; the discordance instead triggers an actionable multimodal safety net comprising (i) serial sectioning, deeper recuts and C4d immunofluorescence or immunohistochemistry on the existing biopsy blocks; (ii) assessment for de novo donor-specific antibodies; (iii) assessment of left ventricular global longitudinal strain by echocardiography and of myocardial oedema by T1/T2 mapping on cardiac magnetic resonance; (iv) verification of calcineurin-inhibitor trough levels and patient adherence; and (v) repeat dd-cfDNA within one to two weeks to establish the biological trajectory of the marker, since a value that continues to rise carries a different implication from one returning toward baseline. Where these signals are concordantly reassuring, the elevation may reasonably be attributed to non-rejection injury and surveillance continued; where they are not, re-biopsy or empiric treatment warrants consideration. This is precisely the decision node that our decision curve analysis validates: the repeat-if-borderline strategy dominates because it resolves ambiguous and discordant molecular signals through serial, multimodal assessment rather than through binary empiric overtreatment or premature discharge. We acknowledge that this scenario, together with the persistently indeterminate result described above, delineates the outer boundary of what a fractional biomarker interpreted against a sampling-limited reference standard can resolve, and that its resolution will require prospective designs in which discordant pairs are followed forward rather than scored as errors at the point of measurement.

Rejection-type specificity. dd-cfDNA thresholds are expected to differ between ACR and AMR given their distinct shedding kinetics as discussed above, and a type-specific analysis would be clinically valuable. This was not possible here: all 14 included studies report rejection as a composite endpoint, and type-stratified 2×2 data are not consistently available in the primary literature and cannot be reconstructed from composite counts. Separate estimation therefore requires individual-patient data, which we identify as the principal objective of the individual-patient-data meta-analysis proposed below. We note that a single composite threshold necessarily averages over two rejection phenotypes with different operating characteristics, itself further evidence that one cutoff cannot serve all diagnostic purposes.

Future work on dd-cf-DNA is crucial, and we need more prospective validation studies using a single, specified assay; we also recommend an individual-patient data meta-analysis to harmonize thresholds across assays. In addition, it is necessary to develop multimodal decision frameworks in the gray/indeterminate zone by integrating other biomarkers (e.g., GEP, DSA) to better inform clinicians; finally, this decision framework can be extended to other solid organ transplants.

## Conclusions

This analysis was conducted to determine whether a single optimal dd-cfDNA threshold exists for post-transplant rejection surveillance and to provide a decision framework. We conclude that there is no single cutoff can simultaneously serve the competing clinical imperatives of screening and confirmation

In our pooled analysis, a result below the rule-out anchor (0.16%) indicated low rejection risk, for which routine surveillance is reasonable; the one-in-five rejection episodes that fall below this anchor are predominantly low-grade, non-cytolytic events, errors of the histological reference standard, or fractionally suppressed signals, and a low result therefore lowers rather than abolishes the probability of rejection. A result above the rule-in anchor (0.48%) warranted strong consideration of biopsy, and where the biopsy is negative, multimodal investigation rather than empiric treatment, since at surveillance prevalence the majority of results above this anchor are not acute rejection; and results in the intermediate zone were genuinely indeterminate, best managed with repeat testing, echocardiography, gene-expression profiling, or donor-specific antibody assessment. Decision-curve analysis demonstrated that no single testing strategy dominates across all clinical contexts; the optimal approach depends on rejection prevalence and the clinician’s tolerance for false positives.

Critically, these anchors are pooled estimates, not universal constants. The deliverable of this work is the tri-state architecture itself, rule out at low dd-cfDNA, rule in at high, and resolve the indeterminate zone before biopsy, not the specific numerical thresholds. We therefore recommend that transplant programs calibrate this framework to their own assay platform and patient population, using their own institutional data in collaboration with their biostatisticians, to derive locally valid rule-out and rule-in anchors. The value of dd-cfDNA lies not in a single number, but in a flexible, context-dependent decision framework that matches the threshold to the clinical question.

## Data Availability Statement

No new data were generated or analyzed in this study. All data included in this analysis are from previously published sources, as cited in the manuscript. All analysis code used in this meta-analysis is publicly available at https://github.com/dr-jabezdavid/dd-cf-dna-analysis under the MIT License.

## Words limit

9,330 words (Introduction to Conclusions, excluding abstract, references, tables and figure legends)

## Ethics Statement

Ethical approval was not required for this study, as it is an analysis of previously published research. All primary studies included in the analysis had obtained approval from their respective Institutional Review Boards (IRBs) or ethics committees, as reported in the original publications. No new data were collected, and no patients or human participants were directly involved in this study.

## Conference Presentation

An earlier version of this abstract was presented at the American College of Cardiology (ACC) 2026 Scientific Session in New Orleans (Abstract ID: 8-ACC, *Optimal Donor-Derived Cell-Free DNA Threshold for Heart Transplant Rejection: A Bayesian Hierarchical Summary ROC Meta-Analysis*). Since that presentation, the work has been substantially revised and expanded for this manuscript. The abstract was published in the *Journal of the American College of Cardiology (JACC)*, Volume 87, Number 13_Supplement.

## Acknowledgments

The authors would like to sincerely thank Dr. Rajesh Menon for his mentorship and valuable advice on the preparation of this manuscript. His guidance greatly contributed to the refinement and clarity of the work.

