## Supplementary material for "Redefining Non-Invasive Post-Transplant Surveillance: A Bayesian Meta-Analysis and Decision Curve Framework for Donor-Derived Cell-Free DNA in Heart Transplantation": Updated Supplementary Material (from another version of this preprint: version 2, 2026-07-06)

Table 1 : Search Strategy

| PUBMED | (((((("heart transplantation"[All Fields] OR ("heart transplantation"[MeSH Terms] OR ("heart"[All Fields] AND "transplantation"[All Fields]) OR "heart transplantation"[All Fields]) OR "heart transplant"[All Fields] OR "transplant recipients"[All Fields] OR "heart transplant recipients"[All Fields] OR "cardiac transplantation"[All Fields] OR "cardiac transplant"[All Fields] OR "transplant rejection"[All Fields] OR "acute rejection"[All Fields]) AND "molecular diagnostic techniques"[All Fields]) OR (("cell free nucleic acids"[MeSH Terms] OR ("cell free"[All Fields] AND "nucleic"[All Fields] AND "acids"[All Fields]) OR "cell free nucleic acids"[All Fields] OR ("cell"[All Fields] AND "free"[All Fields] AND "nucleic"[All Fields] AND "acids"[All Fields]) OR "cell free nucleic acids"[All Fields]) AND ("blood"[MeSH Subheading] OR "blood"[All Fields] OR "blood"[MeSH Terms] OR "bloods"[All Fields] OR "haematology"[All Fields] OR "hematology"[MeSH Terms] OR "hematology"[All Fields] OR "haematoma"[All Fields] OR "hematoma"[MeSH Terms] OR "hematoma"[All Fields] OR "haemorrhage"[All Fields] OR "hemorrhage"[MeSH Terms] OR "hemorrhage"[All Fields] OR "haemorrhages"[All Fields] OR "hemorrhages"[All Fields] OR "haemorrhagic"[All Fields] OR "haemorrhaging"[All Fields] OR "hematologies"[All Fields] OR "haematomas"[All Fields] OR "hematomas"[All Fields] OR "hematoma s"[All Fields] OR "hematomae"[All Fields] OR "hemorrhaged"[All Fields] OR "hemorrhagic"[All Fields] OR "hemorrhagical"[All Fields] OR "hemorrhaging"[All Fields])) OR "dd-cfDNA"[All Fields] OR ("Donor-derived"[All Fields] AND ("cell free nucleic acids"[MeSH Terms] OR ("cell free"[All Fields] AND "nucleic"[All Fields] AND "acids"[All Fields]) OR "cell free nucleic acids"[All Fields] OR ("cell"[All Fields] AND "free"[All Fields] AND "dna"[All Fields]) OR "cell free dna"[All Fields])) OR "biomarkers"[All Fields]) AND ("Endomyocardial"[All Fields] AND ("biopsie"[All Fields] OR "biopsy"[MeSH Terms] OR "biopsy"[All Fields] OR "biopsied"[All Fields] OR "biopsies"[All Fields] OR "biopsy s"[All Fields] OR "biopsying"[All Fields] OR "biopsys"[All Fields] OR "pathology"[MeSH Subheading] OR "pathology"[All Fields]))) OR "emb"[All Fields]) AND "Humans"[MeSH Terms]) NOT ("review"[Publication Type] OR "review literature as topic"[MeSH Terms] OR "review"[All Fields]) | 2016 |
| --- | --- | --- |
|  | ("Heart Transplantation"[MeSH Terms] OR "Transplant Recipients"[MeSH Terms] OR "heart transplant recipients"[All Fields] OR "cardiac transplantation"[All Fields] OR "transplant patients"[All Fields]) AND "acute rejection"[All Fields] AND ("Cell-Free Nucleic Acids"[MeSH Terms] OR "Molecular Diagnostic Techniques"[MeSH Terms] OR "Biomarkers"[MeSH Terms] OR "donor-derived cell-free DNA"[All Fields] OR "dd-cfDNA"[All Fields] OR "cell-free DNA testing"[All Fields] OR "molecular diagnostics"[All Fields] OR "plasma dd-cfDNA"[All Fields]) AND ("Biopsy"[MeSH Terms] OR "endomyocardial biopsy" OR "EMB"[All Fields] OR "biopsy standards"[All Fields] OR "ISHLT"[All Fields]) AND ("Sensitivity and Specificity"[MeSH Terms] OR "diagnostic tests, routine"[MeSH Terms] OR "Odds Ratio"[MeSH Terms] OR "Area Under Curve"[MeSH Terms] OR "diagnostic accuracy"[All Fields] OR "AUC"[All Fields] OR "positive predictive value"[All Fields] OR "negative predictive value"[All Fields]) | 59 |
|  | ddcfDNA"[All Fields] AND "emb"[All Fields] AND ("heart transplantation"[MeSH Terms] OR ("heart"[All Fields] AND "transplantation"[All Fields]) OR "heart transplantation"[All Fields]) | 22 |
| EMBASE | ('heart transplantation'/exp OR 'heart transplant' OR 'transplant recipients' OR 'cardiac transplantation'/exp OR 'cardiac transplant' OR 'transplant rejection'/exp OR 'acute rejection') AND ('molecular diagnostic techniques'/exp OR 'cell free nucleic acids'/exp OR 'donor-derived cell free dna' OR 'dd-cfdna' OR 'biomarkers'/exp) AND ('blood'/exp OR 'hematology'/exp OR 'endomyocardial biopsy'/exp OR 'emb') AND [humans]/lim AND [article]/lim NOT ([review]/lim OR 'editorial'/it OR 'conference abstract'/it) | 500 |
| SCOPUS | (TITLE-ABS-KEY("heart transplantation" OR "cardiac transplantation" OR "heart transplant recipients" OR "transplant rejection" OR "acute rejection")  AND TITLE-ABS-KEY("molecular diagnostic techniques" OR "cell-free nucleic acids" OR "donor-derived cell-free DNA" OR "dd-cfDNA" OR "biomarkers")  AND TITLE-ABS-KEY("endomyocardial biopsy" OR "emb" OR "biopsy" OR "blood"))  AND (LIMIT-TO(DOCTYPE, "ar"))  AND (LIMIT-TO(SUBJAREA, "MEDI"))  AND (LIMIT-TO(LANGUAGE, "English"))  NOT (LIMIT-TO(DOCTYPE, "re") OR LIMIT-TO(DOCTYPE, "ed") OR LIMIT-TO(DOCTYPE, "ab")) | 738 |
| cochrane | "heart transplantation" OR "cardiac transplantation" OR "heart transplant recipients" OR "transplant rejection" OR "acute rejection"  AND "molecular diagnostic techniques" OR "cell-free nucleic acids" OR "donor-derived cell-free DNA" OR "dd-cfDNA" OR "biomarkers"  AND "endomyocardial biopsy" OR "emb" OR "biopsy" OR "blood"  AND "trials"  NOT "review" | 873 |

Tables 2:

|  | Study | RISK OF BIAS | | | | APPLICABILITY CONCERNS | | |
| --- | --- | --- | --- | --- | --- | --- | --- | --- |
|  |  | PATIENT SELECTION | PATIENT SELECTION | INDEX TEST | REFERENCE STANDARD | PATIENT SELECTION | INDEX TEST | REFERENCE STANDARD |
| 1. | Paul J. Kim et al., 2022 (18) | ☺ | ☹ | ? | ☺ | ☺ | ☺ | ☺ |
| 2. | Aditya Mehta et al., 2024 (19) | ☹ | ☹ | ? | ☺ | ☺ | ☺ | ☺ |
| 3. | Conor P. O'Halloran et al., 2024 (20) | ☺ | ☺ | ☺ | ☺ | ☺ | ☺ | ☺ |
| 4. | Marta Jiménez-Blanco et al., 2024 (21) | ☹ | ☺ | ☹ | ☺ | ☺ | ☺ | ☺ |
| 5. | Monica Sorbini et al., 2024 (22) | ☹ | ☺ | ☹ | ☹ | ☺ | ☺ | ☺ |
| 6. | Leor Akabas et al., 2024 (23) | ☺ | ☺ | ☺ | ☹ | ☺ | ☺ | ☺ |
| 7. | Jens Böhmer et al., 2023 (24) | ☺ | ? | ? | ☺ | ☺ | ☺ | ☺ |
| 8. | Shriprasad R. Deshpande et al., 2022 (25) | ? | ? | ? | ☹ | ☺ | ☺ | ☺ |
| 9. | Monica Sorbini et al., 2021 (26) | ☺ | ☺ | ? | ☺ | ☺ | ☺ | ☹ |
| 10. | Franziska Knüttgen et al., 2022 (27) | ☺ | ? | ? | ? | ☺ | ☺ | ☺ |
| 11. | Iwijn De Vlaminck., 2014 (28) | ? | ☺ | ☺ | ☹ | ☺ | ☺ | ☺ |
| 12. | Sean Agbor-Enoh et al., 2021 (29) | ? | ☺ | ☺ | ? | ☺ | ☺ | ☺ |
| 13. | Marc E Richmond et al., 2023 (30) | ? | ☺ | ? | ☹ | ☺ | ☺ | ☺ |
| 14. | Keerthi T Gondi et al., 2021 (31) | ☺ | ☺ | ? | ☺ | ☺ | ☺ | ☺ |

☺ = Low Risk

☹ = High Risk

? = Unclear Risk

Figure 1 : PRISMA Flowchart

**Identification of studies via databases and registers**

Records removed *before screening*:

Duplicate records removed (n = 253)

Records marked as ineligible by automation tools (n = 0)

Records removed for other reasons (n = 0)

Records identified from*:

Databases (n = 4,286)

**Identification**

Records screened

(n = 4,033)

Records excluded**

(n = 4,014)

Reports sought for retrieval

(n = 19)

Reports not retrieved

(n = 0)

**Screening**

Reports excluded:

Conference abstracts, short article (n = 2)

Numerical data not available (n = 2)

Duplicates (n = 1)

Reports assessed for eligibility

(n = 19)

Studies included in review

(n = 14)

Reports of included studies

(n = 14)

**Included**

*Consider, if feasible to do so, reporting the number of records identified from each database or register searched (rather than the total number across all databases/registers).

**If automation tools were used, indicate how many records were excluded by a human and how many were excluded by automation tools.

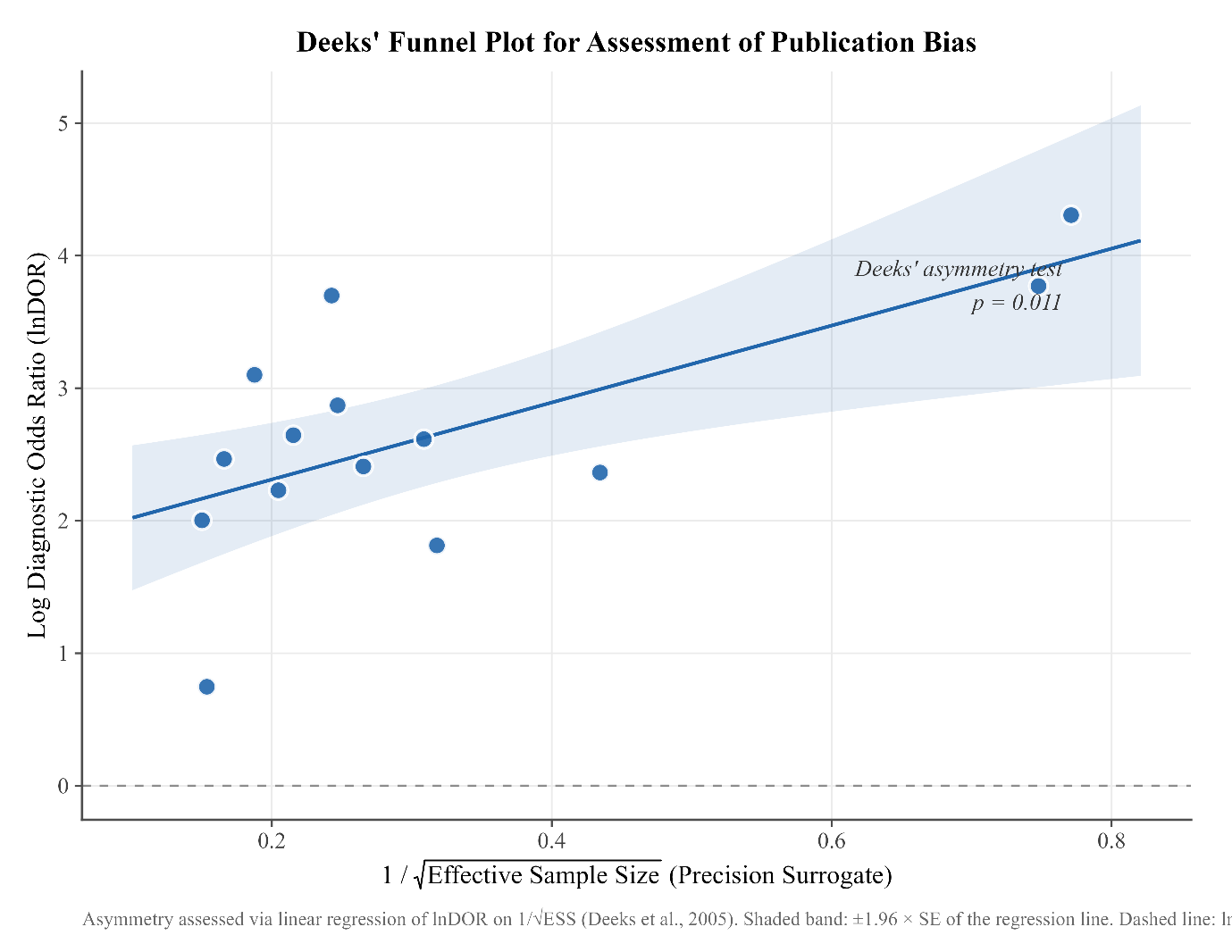

**Figure 2. Deeks' funnel plot asymmetry test for publication bias.**

*Log diagnostic odds ratios from the 14 included studies are plotted against the inverse square root of the effective sample size. A Haldane-Anscombe correction (+0.5) was applied to zero-cell studies. The regression line is shown with a 95% confidence band. A slope of +2.90 (p = 0.0106) indicated statistically significant small-study effects, suggesting that early, smaller studies reported inflated diagnostic accuracy estimates. DD-cfDNA, donor-derived cell-free DNA.*

**Supplementary Methods**

In this section each methodological choice in the manuscript has been presented here as supporting evidence for the logic of the analysis.

**1. Data handling**

No continuity correction was applied to raw cell counts: the JAGS model uses a Binomial likelihood with study‑level random effects, which natively handles zero cells through Beta‑Binomial conjugacy. Adding a +0.5 Haldane Anscombe correction would bias the posterior in a meta‑analysis where several cells are zero by virtue of perfect observed specificity.

The decision threshold is centred at its sample mean (0.208, equivalently 20.8% donor fraction) before entering the regression: *threshold_c[i] = threshold[i] − mean(threshold)*

Centring decorrelates the intercept γ₀ from the slope γ₁, which materially improves MCMC mixing and stabilises posterior summaries on the original threshold scale.

**Table 3.** *Included studies (N = 14)*

| **#** | **Study** | **Tests** | **Patients** |
| --- | --- | --- | --- |
| 1 | Paul et al. | 811 | 223 |
| 2 | Aditya et al. | 384 | 94 |
| 3 | Conor et al. | 101 | 105 |
| 4 | Marta et al. | 1,073 | 206 |
| 5 | Monica et al. (a) | 29 | 29 |
| 6 | Leor et al. | 152 | 152 |
| 7 | Jens et al. | 491 | 52 |
| 8 | Shriprasad et al. | 269 | 269 |
| 9 | Monica et al. (b) | 128 | 19 |
| 10 | Franziska et al. | 293 | 87 |
| 11 | Iwijn et al. | 209 | 65 |
| 12 | Sean et al. | 923 | 165 |
| 13 | Marc et al. | 745 | 144 |
| 14 | Keerthi et al. | 495 | 153 |

*Tests = total number of dd-cfDNA measurements contributed by the study to the meta-analytic pool (TP + FP + FN + TN). Patients = number of unique transplant recipients providing those measurements. The total raw N across all 14 studies is 6,103 measurements from 1,763 patients.*

**2. Cluster‑correction of effective sample size**

Several studies report multiple dd‑cfDNA measurements per patient, violating the independence assumption of the Binomial likelihood. We deflate the study‑level 2×2 cells by a design effect:

$$DEff_{i}=1+(\bar{m}_{i}-1)\text{ }\rho$$

with a primary intra‑patient correlation of ρ = 0.50. Because ρ cannot be estimated from aggregate data, we repeat the entire pipeline for ρ ∈ {0, 0.30, 0.50, 0.70}. The earlier assumption of ρ = 1 (i.e., DEff = m̄) was biologically implausible; the current sweep brackets the empirical range for transplant biomarkers.

**Table 4.** *Per-study cluster characteristics and design effects across the ρ sweep. m̄ = mean cluster size (tests per patient). DEff(ρ) = 1 + (m̄ − 1) · ρ deflates the effective sample size for studies with intra-patient repeated measurements.*

| **Study** | **Tests** | **Patients** | **m̄** | **DEff (ρ=0)** | **DEff (ρ=0.30)** | **DEff (ρ=0.50)** | **DEff (ρ=0.70)** |
| --- | --- | --- | --- | --- | --- | --- | --- |
| Paul et al. | 811 | 223 | 3.64 | 1.00 | 1.79 | 2.32 | 2.85 |
| Aditya et al. | 384 | 94 | 4.09 | 1.00 | 1.93 | 2.54 | 3.16 |
| Conor et al. | 101 | 105 | 0.96 | 1.00 | 0.99 | 0.98 | 0.97 |
| Marta et al. | 1,073 | 206 | 5.21 | 1.00 | 2.26 | 3.10 | 3.95 |
| Monica et al. (a) | 29 | 29 | 1.00 | 1.00 | 1.00 | 1.00 | 1.00 |
| Leor et al. | 152 | 152 | 1.00 | 1.00 | 1.00 | 1.00 | 1.00 |
| Jens et al. | 491 | 52 | 9.44 | 1.00 | 3.53 | 5.22 | 6.91 |
| Shriprasad et al. | 269 | 269 | 1.00 | 1.00 | 1.00 | 1.00 | 1.00 |
| Monica et al. (b) | 128 | 19 | 6.74 | 1.00 | 2.72 | 3.87 | 5.02 |
| Franziska et al. | 293 | 87 | 3.37 | 1.00 | 1.71 | 2.18 | 2.66 |
| Iwijn et al. | 209 | 65 | 3.22 | 1.00 | 1.66 | 2.11 | 2.55 |
| Sean et al. | 923 | 165 | 5.59 | 1.00 | 2.38 | 3.30 | 4.22 |
| Marc et al. | 745 | 144 | 5.17 | 1.00 | 2.25 | 3.09 | 3.92 |
| Keerthi et al. | 495 | 153 | 3.24 | 1.00 | 1.67 | 2.12 | 2.56 |

*DEff < 1 in Conor et al. (m̄ = 0.96) is a numerical artefact of the formula at ρ > 0 when m̄ < 1; the script floors all non-zero cell counts at 1 after deflation, so the effect on the analysis is negligible.*

**Table 7.** *Aggregate effect of the cluster correction across the ρ sweep. The total raw N (6,103 measurements) is deflated to the effective N reported in the analysis.*

| **ρ** | **Mean DEff** | **Median DEff** | **Max DEff** | **Total N (raw)** | **Total N (effective)** | **% retained** |
| --- | --- | --- | --- | --- | --- | --- |
| 0.00 | 1.00 | 1.00 | 1.00 | 6,103 | 6,103 | 100.0% |
| 0.30 | 1.85 | 1.75 | 3.53 | 6,103 | 3,176 | 52.0% |
| 0.50 | 2.42 | 2.25 | 5.22 | 6,103 | 2,518 | 41.3% |
| 0.70 | 2.98 | 2.75 | 6.91 | 6,103 | 2,126 | 34.8% |

*Highlighted row indicates the primary analysis (ρ = 0.50). “Total N effective” is the integer sum of post-deflation cells across all studies and 2×2 categories.*

The stability of conclusions across the sweep (shown later in Table 8) is the strongest argument that the cluster‑correction choice does not drive the findings.

**3. Bayesian HSROC threshold‑regression model**

The conventional HSROC model summarises accuracy at a single implicit threshold. To make the model evaluable at any candidate dd‑cfDNA cut‑off, we include the centred physical threshold as a linear covariate in the positivity‑parameter equation:

$$\lambda_{i}=\gamma_{0}+\gamma_{1}\cdot threshold\_c[i]+u_{i}$$

This allows the posterior predictive SROC curve to be generated across a fine grid, directly mapping the biomarker’s physical scale to diagnostic performance. A quadratic extension (γ₂) was tested to guard against non‑linearity; the 95 % credible interval for γ₂ contained zero in all ρ scenarios, and ΔDIC < 2, supporting the linear specification.

**Hyper‑priors** were chosen to be weakly informative on the logit scale: μθ ∼ N(0, 1), γ₀ ∼ N(0, sd 2), γ₁ ∼ N(0, sd 10), β ∼ N(0, sd 0.5), τθ, τu ∼ Gamma(2, 0.5).

**4. Model evaluation: PSIS‑LOO with exact‑refit fallback**

Leave‑one‑out cross‑validation in a small‑N hierarchical model is challenging because each study carries high leverage. We use Pareto‑smoothed importance sampling (PSIS‑LOO) as a fast approximation, but trigger an exact refit whenever the Pareto‑k diagnostic exceeds 0.7. The exact refit re‑fits the model on N − 1 studies and marginalises analytically over the held‑out study’s random effects with 1,000 inner Monte Carlo draws per hyper‑parameter draw. The final elpd_loo is a blended estimate: PSIS where k ≤ 0.7, exact where k > 0.7.

**Table 5.** *ρ sweep — model fit and influence diagnostics. elpd_loo is the expected log pointwise predictive density; higher (closer to zero) is better. p_loo (blended) = in-sample LPD − elpd_loo and reflects effective model complexity; p_loo (PSIS) is the conventional version from loo::loo(). Pareto-k_max is the worst per-study importance-sampling diagnostic; values above 0.7 trigger exact LOO refits.*

| **ρ** | **elpd_loo (SE)** | **p_loo (PSIS)** | **p_loo (blend.)** | **LOOIC (SE)** | **max Pareto-k** | **# k > 0.7** |
| --- | --- | --- | --- | --- | --- | --- |
| 0.00 | −104.07 (7.03) | 21.06 (1.92) | 32.61 | 185.04 (9.26) | 1.11 | 13 |
| 0.30 | −89.15 (5.28) | 17.91 (1.88) | 24.78 | 164.55 (7.90) | 1.05 | 13 |
| 0.50 | −84.22 (4.89) | 17.08 (1.81) | 22.42 | 157.74 (7.53) | 1.11 | 13 |
| 0.70 | −80.55 (4.81) | 16.13 (1.75) | 20.71 | 151.94 (7.59) | 1.05 | 13 |

*Highlighted row marks the primary analysis. elpd_loo and LOOIC are blended estimates: PSIS where Pareto-k ≤ 0.7, exact-refit where k > 0.7. SE = standard error of the elpd estimate. # k > 0.7 = number of studies that triggered exact refits.*

**Table 9.** *Pareto-k diagnostics and exact-LOO refits at ρ = 0.50. PSIS-LOO flags any study with Pareto-k > 0.7 as having unreliable importance-sampling estimates; the pipeline triggers an exact LOO refit for those studies. At the primary ρ, 13 of 14 studies required exact refits.*

| **Study** | **Required exact LOO refit?** | **Notes** |
| --- | --- | --- |
| Paul et al. | Yes | k > 0.7 at all ρ |
| Aditya et al. | Yes | k > 0.7 at all ρ |
| Conor et al. | Yes | k > 0.7 at all ρ |
| Marta et al. | Yes | k > 0.7 at all ρ |
| Monica et al. (a) | Yes | k > 0.7 at all ρ |
| Leor et al. | Yes | k > 0.7 at all ρ |
| Jens et al. | Yes | k > 0.7 at all ρ; largest deflation |
| Shriprasad et al. | Yes | k > 0.7 at all ρ |
| Monica et al. (b) | No (one ρ only) | Single study with k ≤ 0.7 |
| Franziska et al. | Yes | k > 0.7 at all ρ |
| Iwijn et al. | Yes | k > 0.7 at all ρ |
| Sean et al. | Yes | k > 0.7 at all ρ |
| Marc et al. | Yes | k > 0.7 at all ρ |
| Keerthi et al. | Yes | k > 0.7 at all ρ |

*max Pareto-k at ρ = 0.50 was 1.11; at ρ = 0.30 and ρ = 0.70 it was 1.05. The script's report names the 13 refit studies at each ρ; cross-tabulating across ρ values shows the same study (Monica et al. (b)) consistently has the lowest k, though it is still > 0.7 at three of four ρ values.*

The 13‑of‑14 high Pareto‑k count is **expected** for a 14‑study random‑effects model. It indicates that the fallback was triggered correctly, not that the model is unreliable. The blended elpd_loo combines the gold‑standard exact refits for high‑leverage studies.

**5. Two summary points: marginalised and at‑γ₀**

A single operating point (e.g., the posterior median at the mean observed threshold, γ₀) may mislead readers into thinking the SROC curve can be summarised by one sensitivity/specificity pair. We report two complementary summaries:

- **Marginalised summary:** per posterior draw, sensitivity and specificity are averaged over the empirical distribution of study thresholds. This provides a threshold‑agnostic overall performance estimate.
- **γ₀ point:** the model’s prediction at the mean observed threshold, retained for comparison with traditional meta‑analyses.

In this dataset the two are nearly identical because the threshold distribution is symmetric around its mean, but the distinction is methodologically important.

**6. Adaptive sensitivity and specificity anchors**

**Rationale.** Fixed sensitivity targets (e.g., 95 %) are often unattainable by the posterior and force the analyst to either report an unachievable anchor or impose an arbitrary lower target. Our **adaptive walk‑down** starts at 95 % sensitivity (and 95 % specificity) and lowers the target in 5‑percentage‑point steps until at least 50 % of posterior draws support it. This guarantees that the reported anchors are data‑driven and achievable. The procedure landed on 80 % sensitivity and 90 % specificity at all four ρ values.

**Table 6.** *ρ sweep discrimination, threshold anchors, and operating points. Posterior medians with 95% credible intervals in brackets. The Youden cut-point maximises sens + spec − 1; the sensitivity anchor is the lowest cut-point that delivers ≥ 80% sensitivity; the specificity anchor is the highest cut-point that delivers ≥ 90% specificity.*

| **ρ** | **Pri.** | **AUC** | **Youden cut-point (%)** | **Youden J** | **80%-Sens cut-point (%)** | **90%-Spec cut-point (%)** |
| --- | --- | --- | --- | --- | --- | --- |
| 0.00 |  | 0.784 [0.676–0.841] | 19.42 | 0.539 [0.450–0.620] | 14.61 [5.64–27.74] | 48.89 [35.43–66.83] |
| 0.30 |  | 0.779 [0.671–0.839] | 19.42 | 0.533 [0.437–0.618] | 14.61 [5.64–26.78] | 48.25 [35.75–66.83] |
| 0.50 | ★ | 0.781 [0.668–0.840] | 20.06 | 0.536 [0.437–0.624] | 15.57 [5.96–28.07] | 47.61 [35.11–66.51] |
| 0.70 |  | 0.781 [0.667–0.842] | 20.38 | 0.537 [0.435–0.627] | 15.89 [5.96–29.35] | 47.93 [35.43–66.19] |

*★ marks the primary analysis (ρ = 0.50). All anchor cut-points are reported on the dd-cfDNA % scale. CrI = 95% credible interval.*

**Table 8.** *Range of conclusions across the ρ sweep. min, median, and max are taken across the four sweep values ρ ∈ {0, 0.30, 0.50, 0.70}. This table answers the methodological question: do the headline conclusions change as a function of cluster-correlation choice?*

| **Quantity** | **Min across ρ** | **Median across ρ** | **Max across ρ** |
| --- | --- | --- | --- |
| AUC (median) | 0.779 | 0.781 | 0.784 |
| Youden cut-point (% dd-cfDNA) | 19.42 | 19.74 | 20.38 |
| Youden J (median) | 0.533 | 0.536 | 0.539 |
| Sens-anchor target | 80% | 80% | 80% |
| Sens-anchor cut-point (median, %) | 14.61 | 15.09 | 15.89 |
| Spec-anchor target | 90% | 90% | 90% |
| Spec-anchor cut-point (median, %) | 47.61 | 48.09 | 48.89 |
| elpd_loo | −104.07 | −86.68 | −80.55 |
| p_loo (blended) | 20.71 | 23.60 | 32.61 |
| p_loo (PSIS) | 16.13 | 17.49 | 21.06 |
| max Pareto-k | 1.05 | 1.08 | 1.11 |
| # studies with k > 0.7 | 13 | 13 | 13 |

*All ρ values use the same adaptive sensitivity (80%) and specificity (90%) targets — the targets did not change across the sweep, which means each is achievable in the requisite fraction of posterior draws at every ρ.*

**7. Strategy‑based decision curve analysis**

The DCA evaluates whether a mathematically optimal threshold (Youden) translates into clinical net benefit. To construct joint probabilities for the two sequential‑testing strategies, we use a Gaussian copula on Bernoulli marginals rather than Pearson‑correlation formulas. The Pearson construction imposes a data‑dependent bound on valid correlations; outside that bound the joint probabilities are not legal probabilities. The copula is valid for any ρ ∈ (−1, 1) and any marginal in (0, 1), guaranteeing correct probabilities. Runtime assertions (assert_valid_prob()) verify that every joint probability and every effective sensitivity/specificity lies in [0, 1] for every posterior draw, every threshold probability, and every ρ_repeat value.

**Table 10.** *Decision-curve analysis configuration and validity at ρ_repeat = 0.50. Strategy 5 (repeat-if-borderline) requires the rule-out and rule-in anchors to be appropriately ordered (sens-anchor cut-point < spec-anchor cut-point) at every posterior draw; draws that violate this order are flagged invalid and excluded.*

| **Configuration item** | **Value** |
| --- | --- |
| ρ_cluster (DEff) | 0.50 (primary) |
| ρ_repeat (DCA, default) | 0.50 |
| ρ_repeat sensitivity sweep | 0.30, 0.50, 0.70 |
| Borderline resolution rule (Strategy 5) | Threshold-dependent p_b = 1 − p_t |
| Pre-test prevalence levels | 1-50% |
| Joint-probability construction | Gaussian copula on Bernoulli marginals (pbivnorm) |
| Strategy 5 valid posterior draws | 32,573 / 40,000 (81.4%) |
| — order-inverted draws | 1 |
| — draws with NA anchors | 7,426 |
| Strategy 4 (repeat-if-negative) | 100% valid (no anchor-ordering requirement) |

*The 7,426 NA draws are posterior samples in which either the 80%-sens or the 90%-spec target was unreachable on the threshold grid; these are excluded from the repeat-if-borderline computation. The single order-inverted draw is the count of posterior samples where the sens-anchor cut-point would lie above the spec-anchor cut-point — a sign that the three-zone construction is robust.*

The DCA is repeated across three ρ_repeat values (0.30, 0.50, 0.70) to confirm that the dominance of the repeat‑if‑borderline strategy at low prevalence is robust to uncertainty about the intra‑test correlation.

**8. Software, reproducibility, and computation**

All computation is in R with JAGS via rjags, PSIS‑LOO via loo, and the bivariate normal CDF via pbivnorm. The pipeline is fully reproducible: all random number generation is seeded, MCMC chains are initialised with reproducible seeds, and the entire analysis script is publicly available at <https://github.com/dr-jabezdavid/dd-cf-dna-analysis>
